# Selection and surveillance of 5S ribosomal RNA genes in human populations

**DOI:** 10.64898/2026.08.27.26361558

**Authors:** Laura Sengl, Ivan Bagaric, Clement Conil, Yoann Seeleuthner, Martin B.D. Müller, Simon Mages, Johanna Klughammer, Aurelie Cobat, Jonathan Bohlen

## Abstract

The 5S ribosomal RNA gene is present in the human genome not once but in ∼80 copies, arranged head to tail in a single array of ribosomal DNA on chromosome 1 - one of the most repetitive and least explored regions of the genome. Its product is one of the four RNAs in every ribosome and, when ribosome assembly fails, it activates the tumour suppressor p53. Whether these copies vary in sequence between people, and whether such variation has physiological or pathological consequences, is unknown. Using telomere-to-telomere genome assemblies, whole-genome sequences from ∼490 000 UK Biobank participants, and ∼940 GTEx transcriptomes, we find that every person carries copies bearing substitutions or indels, and that ∼10% of people express such variant 5S rRNA. Mutating every position of the gene *in vitro*, we find that variants blocking incorporation into the ribosome map to the uL5/uL18 interface and activate p53. Remarkably, these same variants are depleted from human populations: selection has acted on the step that p53 monitors. Ribosomal DNA is thus a functional source of human genetic variation, long invisible to genome-wide analysis and shaped by the p53 pathway it controls.

**HIGHLIGHTS:** A single chromosome-1 array makes every person a carrier of variant 5S rRNA genes Saturation mutagenesis maps 5S variants that block assembly and activate p53 Assembly-blocking variants are purged from human populations via p53 surveillance

**eTOC:** Sengl et al. resolve sequence variation across the human 5S rDNA array, one of the genome’s least-explored regions. Variant 5S rRNAs are expressed; and variants that block ribosome assembly activate p53 and are removed from human populations by selection.

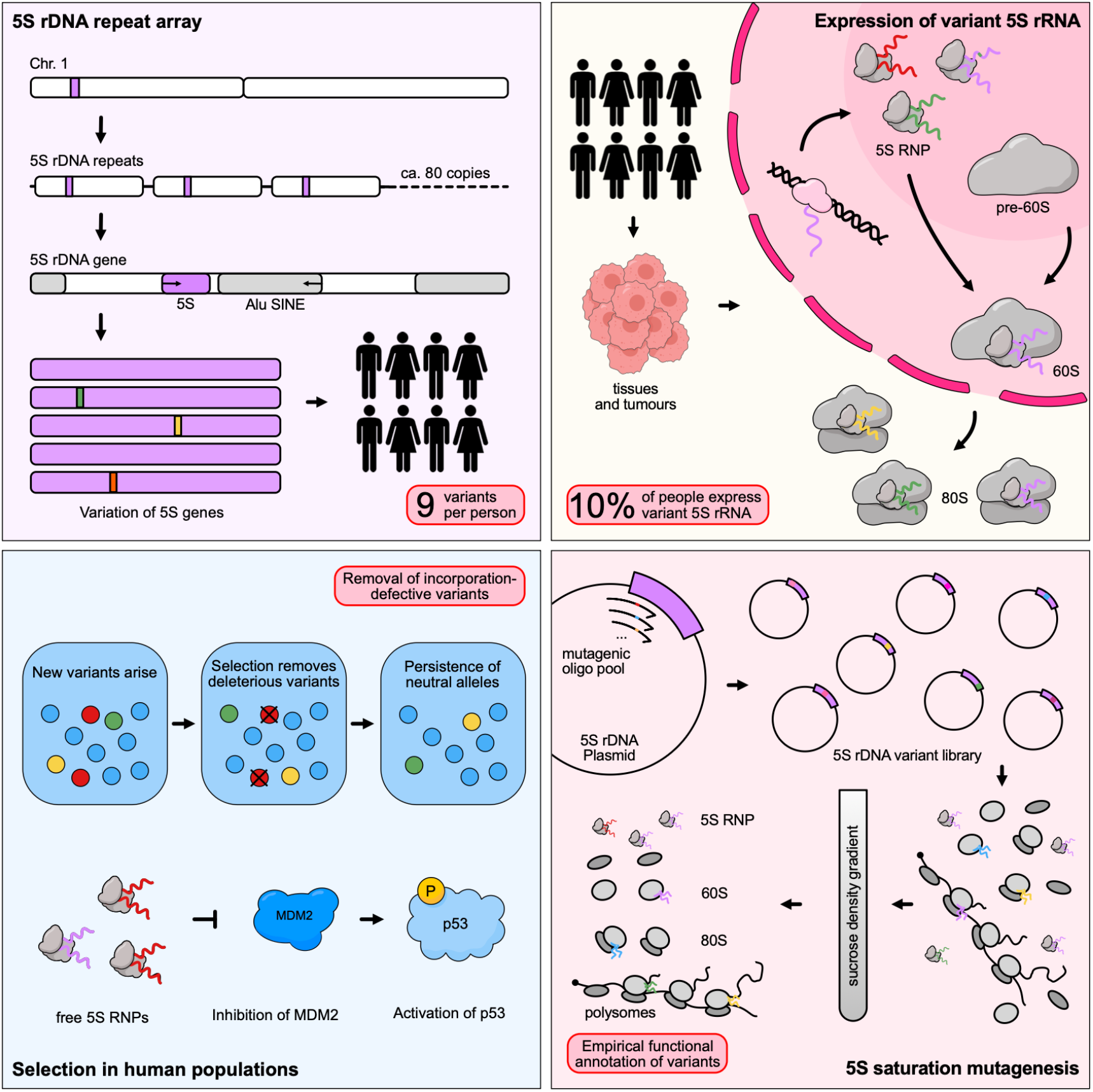

## INTRODUCTION

Ribosomal RNA (rRNA) is the most abundant RNA in the cell and an essential component of every ribosome. To meet this enormous demand, eukaryotes carry not a single copy of each rRNA gene but hundreds, arranged head to tail in tandem ribosomal DNA (rDNA) arrays. Humans have two distinct rDNA classes; the 45S rDNA, encoding the 18S, 5.8S and 28S rRNAs and distributed across the short arms of the five acrocentric chromosomes, transcribed by RNA polymerase I (POLI); and the 5S rDNA, a single large tandem array on chromosome 1q, transcribed independently by RNA polymerase III (POLIII)^1^. Due to their repetitive nature, both arrays long resisted assembly and were largely excluded from genetic analysis until the recent Telomere-to-Telomere reference and long-read sequencing made them tractable^2^.

Both rDNA arrays vary enormously in copy number between individuals, from several hundred 45S copies to ∼80-100 5S copies per array. Despite encoding parts of the same machine, the two arrays copy number varies essentially independently of each other^3–5^. Dosage control of 5S is of special interest because 5S rRNA is more than a structural component of the large ribosomal subunit. With ribosomal proteins uL18 and uL5 it forms the 5S ribonucleoprotein particle (5S RNP), which upon perturbation of ribosome biogenesis accumulates and sequesters MDM2 to stabilize p53^6–8^. The 5S RNP is thus a central surveillance node coupling ribosome assembly to the p53 tumour-suppressor pathway, with direct relevance to ribosomopathies and cancer^9^. It also implies that the 5S array carries regulatory and selective constraints not shared by the 45S: variants that alter 5S expression or its incorporation into ribosomes could perturb this circuit, making the 5S array a candidate target of selection in populations and cancers, where p53 is a central suppressor pathway.

Beyond copy number, rDNA arrays harbor abundant sequence variation between copies and individuals, long proposed to generate heterogeneous ribosomes with specialized functions^10,11^. For the 45S arrays this is beginning to be explored: long-read and population-scale studies have catalogued common and rare rRNA variants, many in the enigmatic surface-exposed expansion segments, defined heritable “ribosome subtypes,” and linked them to human traits and disease^12–14^. Purifying selection on human 45S rDNA genes has been inferred from evolutionary conservation^15–17^. The 5S locus, by contrast, remains largely unexplored: 5S sequence variation has been noted^3^, but whether 5S variants are expressed, if they are incorporated into ribosomes, and their phenotypic consequences remain unknown.

It remains unclear if rDNA variation is functional, and whether such signals are causal or associative. Here we trace human 5S rDNA from genome to ribosome. We catalogue 5S sequence variation across ∼600 telomere-to-telomere haplotypes, ∼490 000 UK Biobank genomes and expression across ∼940 GTEx individuals, then saturation-mutagenise the 5S rRNA gene to measure how each variant is expressed and whether it is incorporated into the assembling ribosome. Reading this functional map against allele frequencies and p53 status in human tumours, we ask which 5S variants the cell expresses, which it incorporates, and which are ultimately removed by selection.

## RESULTS

### The structure of human 5S rDNA arrays from telomere to telomere genomes

To study 5S array structure, we used human T2T assemblies, which now often resolve the array end to end. We collected CHM13 (the first telomere to telomere assembly), Genome in a bottle HG002 (GIAB), Human Pangenome Reference Consortium (HPRC), Chinese Pangenome Consortium (CPC) assemblies, totalling 294 diploid donors (589 haplotype assemblies, only one assembly from homozygous CHM13) (**Table S1**)^2,18^. We then identified the position of the 5S rDNA array in the assembly (**Figure 1A**). The majority (82.7%) of assemblies contained an uninterrupted 5S rDNA array, while 16.8% had the array split into two or three contigs, which can still be unambiguously arranged. Only 0.5% had a fragmented 5S array with more than three contigs, which did not allow unambiguous ordering of the segments. We annotated the number, position and sequence variation of repeat copies within these assemblies. The human 5S rDNA repeat is a 2,168 bp, GC-rich (68%) unit containing a single 120 bp 5S rRNA gene (positions 630–749), transcribed by POLIII from an entirely internal type-1 promoter, with transcription terminated by an oligo-dT tract immediately 3′ of the gene (750–765). The cis-regulatory footprint is therefore compact (∼140 bp) and proximal, leaving ∼94% of the unit as non-transcribed spacer (NTS). The 5′ spacer (1–629) is G-rich and bounded by a CpG/(GT)_n_ low-complexity junction, while the larger 3′ spacer (766–2,168) is C-rich and contains a single, near-full-length AluY-related SINE inserted antisense (787–1,066; ∼93% identity) and terminates in a long pyrimidine (CTGT/CTCT)_n_ microsatellite (2,009–2,155) (**Figure 1A**, **Table S2**). We built assembly maps for the 589 haplotypes which annotate the single nucleotide variants (SNVs) and insertion-deletion variants (indels) in each of the copies (**Table S3**). The HG002 maternal-haplotype map (104 copies) shows scattered private variants and several multi-copy variant tracts, where neighboring copies, sometimes for large stretches carried the same substitution (**Figure 1B**). To make these data accessible, the complete set of 5S rDNA variants, their population frequencies, secondary-structure context, and functional scores can be explored interactively at https://genesintranslation.com/. In summary, human T2T genome assemblies reveal the architecture and variation of the 5S rDNA tandem repeat array.

**Figure 1.**
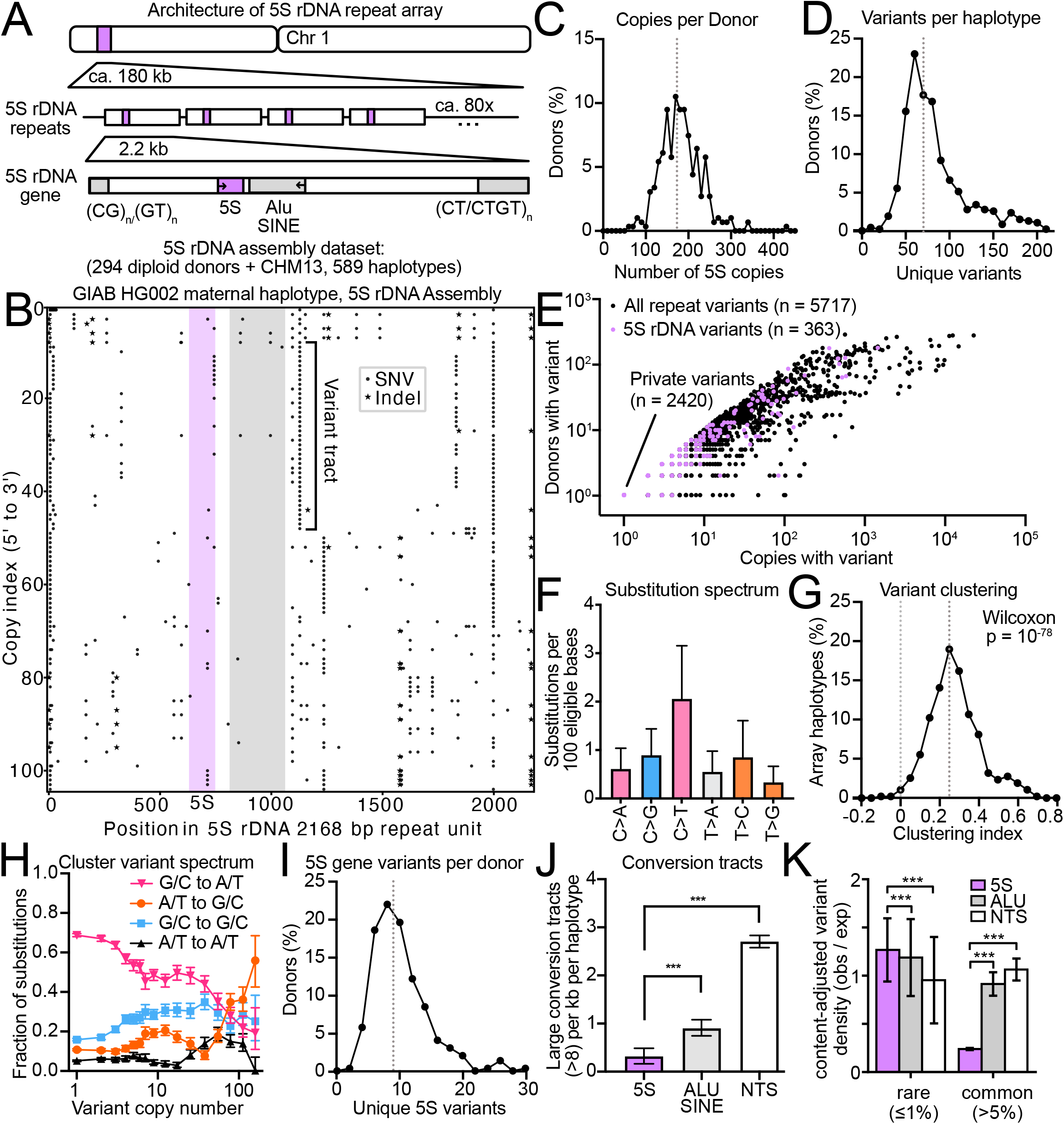
The human 5S rDNA array carries abundant, gene-conversion–shaped sequence variation. **(A)** Architecture of the 5S rDNA locus. A single ∼220 kb tandem array on chromosome 1 contains ∼80 copies of a 2.2 kb repeat unit; each unit comprises the 120 bp 5S rRNA gene, an antisense Alu SINE, and flanking GC-rich microsatellites [(CG)n(GT)n at the 5′ junction, (CT/CTGT)n at the 3′ end]. The assembly variant catalog combines CHM13, GIAB HG002, HPRC, and Chinese Pangenome Consortium (CPC, East-Asian) genomes (294 donors + CHM13, 589 haplotypes). **(B)** Per-copy variant map of the GIAB HG002 maternal haplotype: each row is one repeat copy (ordered 5′→3′ along the array), each mark a variant relative to the population consensus unit (dots, SNVs; stars, indels). The 5S gene and antisense Alu are shaded; a representative contiguous variant tract is bracketed. **(C)** Distribution of haploid 5S copy number across donors; red dashed line, median. **(D)** Distribution of unique variants (SNVs + indels) per haplotype; red dashed line, median. **(E)** Per-variant prevalence: number of array copies versus number of donors carrying each variant. 5S rDNA gene variants (red, n=363) are shown over all repeat-unit variants (black, n=5,717); 2,420 variants are private (single copy). **(F)** Genome-unit substitution spectrum, expressed as substitutions per 100 eligible bases for each of the six mutation classes (C>A, C>G, C>T, T>A, T>C, T>G). Bars, mean ± SD across haplotypes; C>T predominates. **(G)** Within-array clustering of variants. Distribution of the per-haplotype clustering index; values >0 indicate that variants are spatially clustered along the array. Observed versus position-scrambled, Wilcoxon p=10^-78^. **(H)** Substitution spectrum of clustered variants as a function of variant copy number. Fraction of each directional class [A/T→G/C, G/C→G/C, A/T→A/T, G/C→A/T]; the rising A/T→G/C fraction with tract size is the signature of GC-biased gene conversion. Points, fraction; bands, Wilson 95% CI. **(I)** Distribution of unique 5S gene-region variants per donor; red dashed line, median. **(J)** Rate of large gene-conversion tracts (>8 contiguous carrier copies) per kb per haplotype, by region (5S gene, Alu SINE, other NTS). Bars, mean ± 95% CI; the 5S gene is specifically depleted of large tracts. significance by paired Wilcoxon signed-rank test between regions (overall Friedman test across regions). ***p<0.001. **(K)** Content-adjusted density of rare (<=1% of haplotypes) and common (>5%) single-nucleotide variants, by region, 5S gene: purple; Alu SINE: grey, other NTS spacer: white, bars, mean +/- 95% CI across 589 haplotypes. Significance by paired Wilcoxon signed-rank test (5S vs other NTS): ***p < 0.001.

### 5S rDNA repeat array variation is driven by GC-biased gene conversion

Among the 5S rDNA array donors and haplotypes studied, copy number varied up to 7-fold (**Figure 1C, S1A)**, raising the question of how dosage control is achieved amid this vast copy number variation. 5S rDNA arrays were rich in sequence variation, harboring a median number of around 60 unique variants per array (**Figure 1D**), with many unique variants being present in multiple copies of the same haplotype or donor (**Figure 1E**). Copy number correlated well with number of unique variants, generating more diversity in bigger arrays (**Figure S1B**). We mapped the six-class substitution spectrum of the repeat unit, collapsing substitutions by strand symmetry and found that cytosine deamination products (C>T) were the dominant type of substitution in the array (**Figure 1F**). We next investigated whether the variant tracts, observed for example in copies 8-52 at position 1100 of HG002 mat (**Figure 1B**), were a general feature of 5S rDNA arrays. Variant clustering as quantified with a clustering score (score <0: variants regularly spaced, score 0: identical variants are randomly distributed, score> 0: variants form tracts), revealed that variants are significantly clustered along the array (**Figure 1G**). We hypothesized that these variant tracts may arise by non-allelic gene conversion, a mechanism of concerted evolution in which homologous copies are used as repair templates for copies that have acquired damage. One type of gene conversion implicated at rDNA loci^19^ is biased towards A/T to G/C conversion (gBGC) and mediated by the mismatch repair machinery^20^. Higher copy number variants were increasingly G/C-biased (**Figure 1H**), suggesting variant tracts and high copy number variants in 5S rDNA arise by non-allelic gBGC. Surprisingly, within the highly conserved 5S gene itself, each donor carried a median of nine distinct variants (unique substitutions, each of which may recur across several array copies, **Figure 1I**). Whether these variants are expressed or affect physiology is not known. We asked whether the gene is subject to purifying selection relative to the rest of the repeat unit. Indeed, we observed that the 5S gene was the least likely to contain large gene conversion variant tracts compared to the ALU SINE or NTS (**Figure 1J**). Furthermore, when an array contained a large number (>8) of copies with the same variant, these were less likely to be clustered when in the 5S gene (**Figure S1C**). As a result, common variants were strongly depleted from the 5S gene, while rare variants were moderately in excess (**Figure 1K**). In summary, gene conversion is a major driver of 5S rDNA repeat array variation and the transcribed 5S rDNA itself is partially protected from this by evolutionary constraint.

### 5S rDNA variant calling from short-read whole-genome sequencing data

Since the 5S rDNA gene was subject to purifying selection, we hypothesized that the presence of 5S rDNA variation could have physiologically deleterious consequences. We aimed to identify 5S rDNA variants in a clinically annotated cohort. For this, we first built and calibrated a variant calling method on short-read whole-genome sequencing (WGS), as T2T assemblies or even long-read sequencing are typically not available for such cohorts. We used the T2T assemblies and short-read WGS from the HPRC year 1 release to build this variant caller based on an assembly ground-truth. There are numerous 5S rDNA pseudogenes scattered along the genome, so we checked where true 5S rDNA reads would typically map. Simulating short-reads from the transcribed 5S gene copies of the CHM13 T2T assembly and from all 337 dispersed 5S pseudogene loci, then aligning them to GRCh38, we found that array gene reads mapped exclusively to the chr1q42 array and never onto a pseudogene while only 1 in 400 000 pseudogene reads reached the array (extraction precision 99.9997%; **Figure S2A**). Cross-mapping remained zero in both directions across the realistic load of 0 to 2 substitutions per read and stayed below 0.1% even at an extreme 12 substitutions per 150 bp read (∼8% divergence; **Figure S2B**). Short-reads cannot be assigned to individual array copies within chr1q42: they map ambiguously across the repeat, precluding per-copy variant calling. To resolve this, we extracted all reads mapped to the repeat array and re-aligned them to a single, population consensus of the 5S rDNA repeat, producing an even, ca. 4800x coverage along the repeat unit (**Figure S2C**). We then called variants allowing for heterogeneous variant allele frequencies (VAF), given that a single variant copy in a typical individual (ca. 80 copies per Chromosome 1) is expected to give VAF = 0.62%. Although this method showed good concordance with the assembly ground truth (**Figure 2A**), it struggled to identify single-copy variants (65% sensitivity versus 82% overall sensitivity, **Figure S2D**) and produced a high false-positive rate at low VAF likely due to sequencing errors (**Figure S2E**). We reasoned that assemblies may sometimes miss single-copy variants and therefore aimed to investigate the false positive variant calls further. We reasoned that some apparent false positives might be real variants missed by the assembly, and tested this against orthogonal PacBio HiFi data (HPRC). Indeed, 45% of false positives were also detected in PacBio sequencing, a significant overlap (p<5×10⁻⁴), because HiFi chemistry is orthogonal, shared errors are unlikely, so these are genuine variants the assembly missed. At a 0.3% VAF threshold the caller reached 82% sensitivity at 69% precision (**Figure 2C**), the optimum from a threshold sweep of the rescued calls. In short, short-read WGS calls 5S rDNA variants with accuracy sufficient for population-scale variant discovery and frequency estimation, consistent with established short-read analyses of 45S rDNA sequence variation in population cohorts^12,21^, though below what per-individual clinical genotyping requires.

**Figure 2.**
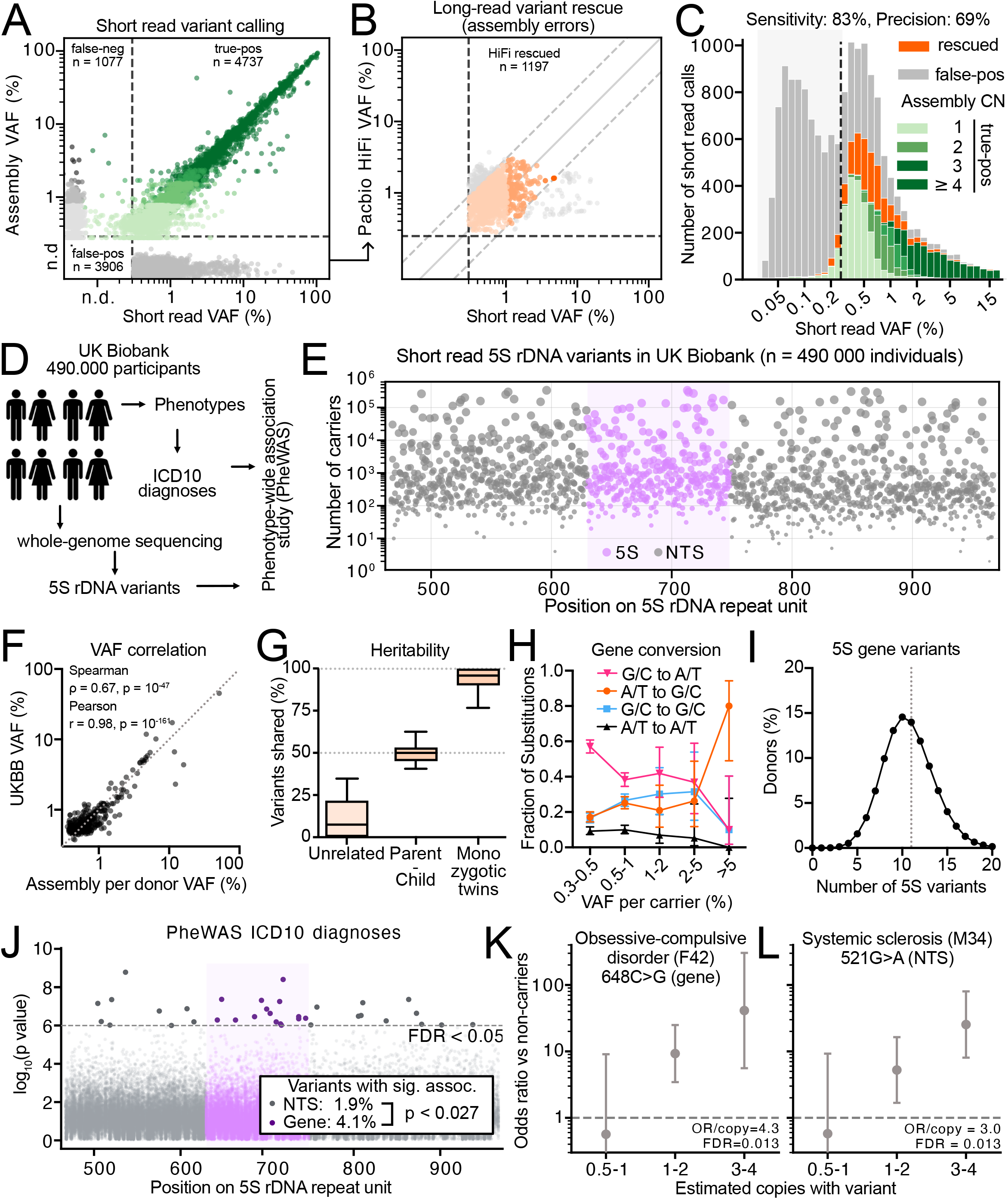
Short-read genotyping of 5S rDNA variation across 490 000 UK Biobank participants enables population and phenome-wide analysis. **(A)** Benchmarking of short-read variant calling against matched genome assemblies. Per-variant short-read VAF versus assembly VAF; true positives (n=4,737), false positives (not detected in assembly, n=3,906), and false negatives (n=1,077) are indicated. n.d., not detected. **(B)** Long-read rescue of apparent false positives. PacBio HiFi VAF versus short-read VAF for calls absent from the assembly; 1,197 are confirmed by HiFi and thus reflect assembly collapse rather than calling error. **(C)** Short-read call composition versus VAF, as a stacked histogram of false positives (gray), HiFi-rescued calls (orange), and true positives colored by assembly copy number (1, 2, ≥3). At the 0.3% calling threshold (dashed), overall sensitivity is 81.5% and precision 69.2%. **(D)** Schematic: UK Biobank whole-genome sequences (∼490 000 individuals) yield 5S rDNA variants and ICD-10 diagnoses for a phenome-wide association study (PheWAS). **(E)** Population carrier landscape along the 5S repeat unit: number of carriers per variant versus position, 5S gene region shaded. Each point is a variant. **(F)** Concordance of population and assembly allele frequencies on a matched per-individual basis: UK Biobank within-person VAF versus assembly per-donor (diploid) VAF (all HPRC, 233 donors; n=356 variants). Both are the mean over confident carriers (VAF≥0.3%), with assembly VAF computed per donor by pooling both haplotypes’ copies so the denominators match. Spearman ρ=0.67 (p=3.4×10^-47); Pearson r=0.98 (p=6.7×10^-161). **(G)** Heritability of 5S variation: percentage of variants shared between unrelated individuals, parent–child pairs, and monozygotic twins (box plots; median, IQR, whiskers). **(H)** Directional substitution spectrum as a function of within-carrier VAF, showing enrichment of A/T→G/C (gene conversion / gBGC) at higher VAF. Points, fraction; bars, 95% CI. **(I)** Distribution of the number of distinct 5S gene variants per UK Biobank carrier (VAF≥1%); every individual carries ≥1 (mean 10.7, median 11). Red dashed line, median. **(J)** PheWAS of 5S variants against ICD-10 diagnoses: -log_10_(p value) versus position on the repeat unit. Gene-region variants (shaded) are enriched for FDR<0.05 associations relative to the surrounding NTS (4.1% vs 1.9%; OR=2.2, permutation p=0.027). Red dashed line, FDR=0.05. **(K)** Dosage– response for a representative gene association (obsessive-compulsive disorder, F42; 648C>G): odds ratio versus non-carriers across estimated variant-copy bins. Points, OR; bars, 95% CI; OR/copy=4.3, FDR=0.013. **(L)** Dosage–response for a representative NTS association (systemic sclerosis, M34; 521G>A): as in **(K)**; OR/copy=3.0, FDR=0.013.

### Prevalent 5S rDNA variation associates with clinical phenotypes

Using this methodology, we called 5S rDNA variants in the region of the array repeat that contains the 5S rDNA transcribed gene and around 200 bp of flanking sequence in the ca. 490 000 probands of the United Kingdom Biobank (UKBB, **Figure 2D, E**). Coverage was homogeneous in this region and averaged around 3000x (**Figure S2F**), consistent with roughly 200 5S rDNA copies per donor. VAFs correlated well between the UKBB and the T2T assemblies despite originating from non-overlapping cohorts with different ethnicity composition (**Figure 2F**). Variants were consistently inherited within families (**Figure S2G**), for example parent-child pairs (n = 3000) shared around 50% of their variants, monozygotic twins (n = 50) shared essentially all their variants (>90%), while randomly paired people shared only a minority of variants (ca. 10%, **Figure 2G)**. This validated that most detected variants are true positives. Furthermore, variants that occurred at higher VAFs, which are more likely to occur in variant tracts, exhibited a substitution bias indicative of gBGC (**Figure 2H**), further confirming the consistency with ground truth variants from T2T assemblies. Similar to T2T assemblies, UKBB probands harboured a median of eleven unique 5S gene variants (**Figure 2I**). Next, we performed a phenotype-wide association study (PheWAS) analysis on International Classification of Diseases revision 10 (ICD-10) diagnoses, modelling each variant’s per-proband copy number, estimated by VAF, as a quantitative predictor in covariate-adjusted logistic regressions (age, sex, 10 genetic principal components). Thus, hits reflect dose-dependent effects. We observed 34 significant phenotype-genotype associations, which were enriched in the gene body (p<0.03, **Figure 2J, Table S4**), consistent with changes inside the transcribed gene being more likely to contribute to disease. Finally, we show two such dose-dependent enrichments for obsessive-compulsive disorder (OCD, **Figure 2K**) and systemic sclerosis (**Figure 2L**) driven by association to variants in the gene and NTS, respectively. In summary, variation in the 5S rDNA repeat array, in particular in the transcribed 5S rRNA gene associates with risk for clinical disease in a dose-dependent manner.

### Detection of expressed 5S rDNA variants in GTEx cohort

We asked whether sequence variants in the 5S rDNA gene may be expressed in cells and tissues of their carriers. We utilized the GTEx cohort which provides WGS and tissue-resolved bulk RNA-seq of 943 organ donors (**Figure 3A**). We called 5S rDNA variants from the short-read WGS data as described above. Next, we hypothesized that it would be possible to detect residual reads on ribosomal RNA from the poly-A enriched bulk RNA-seq datasets. We filtered for read-pairs in which both reads mapped exclusively inside the 5S rRNA sequence to eliminate potential contamination from genomic DNA. With this approach, we were able to achieve a median coverage of ca. 900x per donor (**Figure 3B**). We then called variants on these rRNA reads and compared RNA VAF in groups of carriers and non-carriers for all genetically present 5S rDNA variants. This revealed 52 out of 160 genetic variants with statistically significant levels of expression compared to non-carrier background (**Figure 3C, D, Table S5**). 5S rRNA is not polyadenylated and is therefore captured only as residual carryover in poly-A libraries, this limits sensitivity, not specificity: carriers and non-carriers are assayed identically, so any technical or genomic-DNA background is shared between groups, and a variant-allele excess in carriers reflects genuine expression rather than capture bias. Selection of read pairs that strictly cover only the 120 nt of rRNA and not genomic context excludes the possibility of cross-contamination from gDNA. Expressed variants were mildly enriched in the 3’region of the 5S rRNA (**Figure 3E**). Around 10% of GTEx probands detectably expressed at least one 5S rRNA variant (**Figure 3F**). RNA and DNA VAF correlated weakly overall (ρ=0.1) but strongly at individual loci (e.g. 687G, ρ=0.82; **Figure 3G**). RNA VAF was ∼7-fold below DNA VAF, indicating that variant expression is suppressed relative to genomic dosage. Finally, we investigated whether specific tissues or organs may be more likely to express 5S rRNA variants. We found 5S rRNA variant expression to be highest in brain (**Figure 3H**), consistent with the elevated expression of 45S variants in the brain of mice^3^. In summary, around 10% of people detectably express variant 5S rRNA, potentially contributing to structural ribosome heterogeneity and functional impairment of the translational machinery.

**Figure 3.**
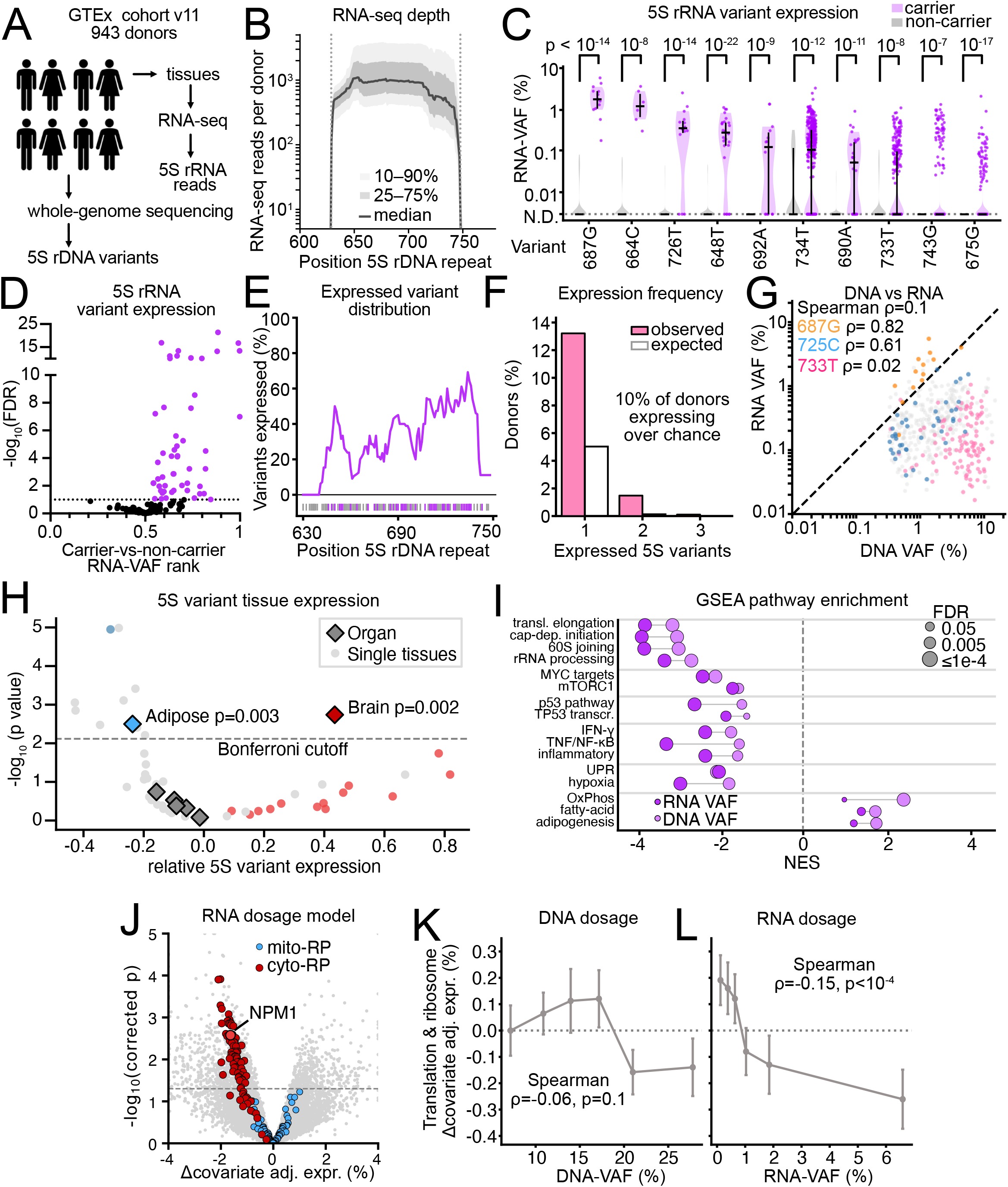
A substantial fraction of humans express variant 5S rRNAs that dose-dependently downshift expression of ribosomal protein genes. **(A)** Schematic: GTEx donors (whole-genome sequencing → 5S rDNA variants; bulk RNA-seq → pooled-per-donor 5S rRNA reads) are used to test variant expression (943 donors). **(B)** Per-donor pooled RNA-seq depth along the 5S repeat (median; bands, q25–q75 and q10–q90). **(C)** Per-variant carrier-versus-non-carrier RNA-VAF for a curated 10-variant set ordered by carrier median (truncated violins; carrier, red; non-carrier, gray). Carriers express the variant allele in RNA (Mann–Whitney U, Benjamini-Hochberg; all q≤3×10^-6). N.D., not detected. **(D)** Threshold-free rank-skew volcano: per-variant carrier-vs-non-carrier RNA-VAF rank statistic (AUC) versus -log10 FDR for 160 testable variants. 52 are carrier-HIGH (expressed); none show higher expression in non-carriers. **(E)** Positional distribution of expressed variants: sliding-window fraction of carrier-HIGH variants along the gene; expression concentrates in the 3′ half (peak ∼87% near position 729, where coverage is lower, excluding a depth artifact). Ticks above X-axis: detected (purple) and undetected (gray) variants. **(F)** Prevalence of variant 5S rRNA expression: number of expressed variants per donor, observed (red) versus chance-null (gray). 15% of donors express ≥1 variant (95% CI 34.5–40.4) against a 5% chance rate (∼10% genotype-attributable). Denominator, all 943 donors. **(G)** DNA-VAF versus RNA-VAF at genuine loci (n=757): RNA-VAF is ∼7× lower than DNA-VAF, with locus-specific transcription efficiencies (per-variant Spearman ρ shown for 687G, 725C, 733T). **(H)** 5S-variant tissue expression: per-tissue relative 5S-variant RNA expression versus -log10 p (light circles, individual tissues; dark diamonds, organ-level groups); brain (p=0.002) and adipose (p=0.003) express variant 5S most and least strongly (dashed line, Bonferroni threshold). **(I)** Gene-set enrichment (GSEA) of the meta-analysed trans-effect against 5S dose (RNA dose, red; DNA dose, blue); NES per pathway, dot size ∝ FDR. Translation/ribosome, growth (MYC/mTORC1) and p53 pathways are coordinately downregulated, with DNA and RNA dosage broadly concordant. **(J)** Trans-effect volcano (RNA-dosage model): change in covariate-adjusted expression (%) versus -log10(corrected p); cyto-RP (red) coordinately downregulated, mito-RP (blue) not; *NPM1* labeled. **(K)** Dose-response of the translation/ribosome module versus DNA-VAF (%) (Spearman ρ=-0.06, p=0.1). **(L)** Dose-response versus RNA-VAF (%) (Spearman ρ=-0.15, p<10⁻⁴); the module tracks RNA (expression) dose more than genomic dose.

### Expression of variant 5S reduces expression of ribosomal protein mRNAs

We next asked what gene-expression consequences follow from carrying or expressing variant 5S rRNA. We tested each variant’s transcriptional consequence in a covariate-adjusted, per-tissue differential-expression framework. The covariate correction is essential here: the 19,244 samples (37 tissues, 943 donors) are structured by RNA integrity, ischemic time, age, sex, ancestry and batch. For each donor we defined two continuous, aggregate dosage metrics over all 5S gene-region variants: a DNA dosage, the summed genomic variant-allele fraction across carried variants (from WGS), and an RNA dosage, the summed RNA-seq variant-allele fraction in excess of the non-carrier background. Within each tissue we fit a negative-binomial model (DESeq2), testing the RNA and DNA dosage coefficients individually and in a joint two-predictor model, then combined per-gene effects across the 37 tissues by meta-analysis with permutation correction. The most strongly affected pathways were largely consistent between DNA and RNA VAF and included downregulation of ribosome and mRNA translation, growth signalling and p53 signalling (GSEA, **Figure 3I, Table S6**). All cytosolic ribosomal protein mRNAs (cyto-RP), but not mitochondrial RPs, were downregulated (**Figure 3J, S3A, Table S7**), consistent with a compensatory response to dysfunctional variant 5S rRNA. NPM1, a ribosome biogenesis factor that chaperones components of the 5S RNP^22–24^, was significantly affected too. Other ribosome biogenesis factors were not strongly affected. The effects were modest (1–2% reduction), matching the 1–2% RNA VAFs that drive them, and place this variation within the range compatible with normal physiology: GTEx donors carry it without overt disease. Expressed variants drive this effect; in the joint model the dose-dependent effects of RNA VAF were more consistent and stronger than those of DNA VAF (**Figure 3K, L**). As genetic ancestry is a common confounder, we repeated the analysis in the largest group (white European), obtaining largely congruent results (**Figure S3B, C**). In summary, expressing variant 5S rRNA coordinately downregulates ribosomal protein mRNAs.

### Functional testing of 5S rRNA variants by saturation mutagenesis

We aimed to understand the functional impact of 5S rRNA sequence variation on stability and ribosome incorporation of the molecule. To this end, we cloned one repeat of the 5S rDNA array into a plasmid (**Figure S4A**). Transfection of this plasmid into HEK293 cells led to a significant, ca. 40% upregulation of 5S but not 5.8S rRNA (**Figure S4B, C**), confirming our ability to express 5S rRNA from a plasmid. Next, we performed saturation mutagenesis of this plasmid, successfully generating a library covering 84% of all possible single nucleotide variants in the transcribed region of 5S rRNA (**Figure 4A**). We transfected this plasmid library into HEK293 cells and fractionated the cells using sucrose density centrifugation. We obtained and sequenced 5S rRNA from total input RNA, and fractionated 60S ribosomal subunits, 80S ribosome complexes and polysomes (**Figure 4B**). Variant analysis revealed robust expression of variant 5S rRNA in total 5S rRNA, when compared to total 5S rRNA from mock (GFP) transfected cells (**Figure 4C**), validating the expression of the saturation mutagenesis library in cells. Strikingly, variant 5S rRNA was progressively depleted from 60S, 80S and polysome fractions, indicating exclusion of dysfunctional variants during assembly (**Figure 4C**). In summary, we established a method to investigate the functional impact of 5S rRNA variants at scale.

**Figure 4.**
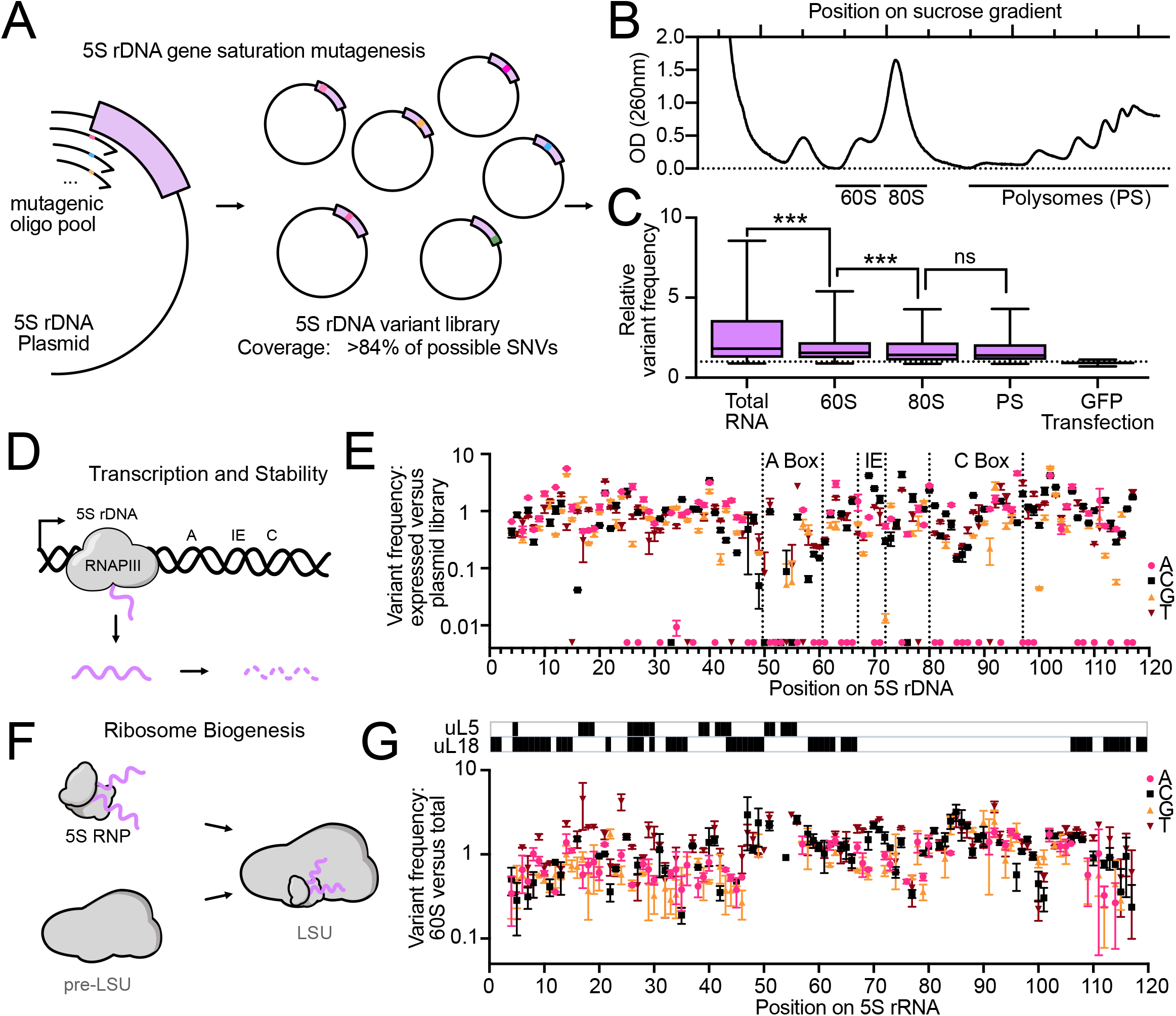
Saturation mutagenesis maps incorporation-defective 5S variants. **(A)** Saturation-mutagenesis strategy: an oligo pool mutagenizes the cloned 5S rDNA gene to build a plasmid variant library covering >84% of possible SNVs. **(B)** Sucrose-gradient absorbance (OD 260 nm) profile of transfected and fractionated cells, resolving free 60S, 80S monosomes and polysomes (PS). **(C)** Relative variant frequency across gradient fractions (total RNA, 60S, 80S, polysomes) and a GFP control (box plots; median, IQR, whiskers); frequency drops from total RNA→60S and 60S→80S (*) with no further change into polysomes (n.s.), localizing selection to 5S incorporation into the large subunit. **(D)** Schematic of the transcription/stability filter (5S-gene internal control elements Box A, IE, Box C). **(E)** Per-position variant frequency in expressed cellular RNA versus the input plasmid library along the 5S gene (nucleotides coloured A/C/G/T; Box A, IE, Box C marked); depletion marks transcription/stability-critical positions. **(F)** Schematic of the ribosome-biogenesis filter: the 5S RNP joins the pre-large subunit (pre-LSU) to form the mature LSU. **(G)** Per-position variant frequency in the 60S fraction versus total RNA along the 5S rRNA (A/C/G/T; uL5/uL18 contact positions marked above); depletion marks incorporation-critical positions.

### Distinct patterns of expression and incorporation perturbation by 5S variants

We then analyzed differences in variant proportion between DNA plasmid library and total expressed 5S rRNA to reveal variants that affected 5S synthesis or stability (**Figure 4D, E**). This revealed large heterogeneity in the expression efficiency of 5S variants over roughly three orders of magnitude (**Figure 4E, Table S8**). Numerous variants that were present in the DNA library were not detected in RNA at all. Reduced or absent expression was most frequent in the internal promoter^25^, most strikingly in the A Box element and also notable in the C Box (**Figure S4D**), confirming the validity of our functional data. Next, we assessed the frequency of 5S variants in total versus 60S incorporated 5S rRNA (**Figure 4F, G**). This assesses the 5S RNP incorporation step that signals towards and activates p53 when defective. Variants that were not detected in total RNA could not be assessed. Variants varied roughly 10-fold in their incorporation efficiency (**Figure 4G**). Incorporation-defective variants clustered in the first 45 nt, coincident with the uL5/uL18 contact footprint (**Figure 4G, S4E**, Fisher OR=3.26, p<0.001). We calculated the change in folding energies of the variant 5S sequences compared to wildtype using RNAfold (**Figure S4F**). Variants that negatively impacted expression were more likely to be destabilizing and variants that negatively impacted incorporation were more likely to be stabilizing (**Figure S4G, H**). In summary, saturation mutagenesis and pooled variant assessment reveals the functional landscape of 5S rRNA variants.

### 5S incorporation defects reduce ribosomal protein mRNA expression

We next tested whether 5S variants that impair 5S expression or incorporation are associated with distinct gene-expression changes in GTEx. To this goal, we grouped the variants according to their experimentally measured functional effects. Here we defined dosage as each donor’s genomic 5S variant load, not as the amount of variant 5S rRNA they express. The reason is circularity: the variant classes we are comparing differ in how well they are expressed, so an expression-based measure of dosage would be depressed by the very defect we are testing for. Genomic load is fixed at the DNA level, independent of a variant’s expression, and thus measures dosage without this circularity. For every variant carried by >10 donors we ran the covariate-adjusted model and meta-analysed across tissues as above, then quantified the effect of copy load on the cytosolic ribosomal-protein (cyto-RP) module. Copy load of incorporation-defective variants (expressed but poorly assembled) coordinately repressed the cyto-RP module (83% of cyto-RP genes downregulated, **Figure 5A**), whereas copy load of fully functional variants did the opposite, moderately raising it (**Figure 5B**). The 5S rDNA genomic dosage thus produces opposite transcriptional consequences depending on whether the encoded 5S can be incorporated. The more a variant was expressed yet failed to incorporate, the more strongly it suppressed the biogenesis program: the strength of cyto-RP repression scaled continuously with a variant’s position on the incorporation–expression axis (ρ=-0.43, p=0.015; **Figure 5C**). Decomposing the axis attributed this largely to incorporation (ρ=+0.36, p=0.044), not expression (ρ=-0.11, p=0.445; **Figure S5A, B**), the same assembly step that is under populational constraint. In summary, genomic load of incorporation defective 5S rRNA variants drives suppression of ribosome mRNAs.

**Figure 5.**
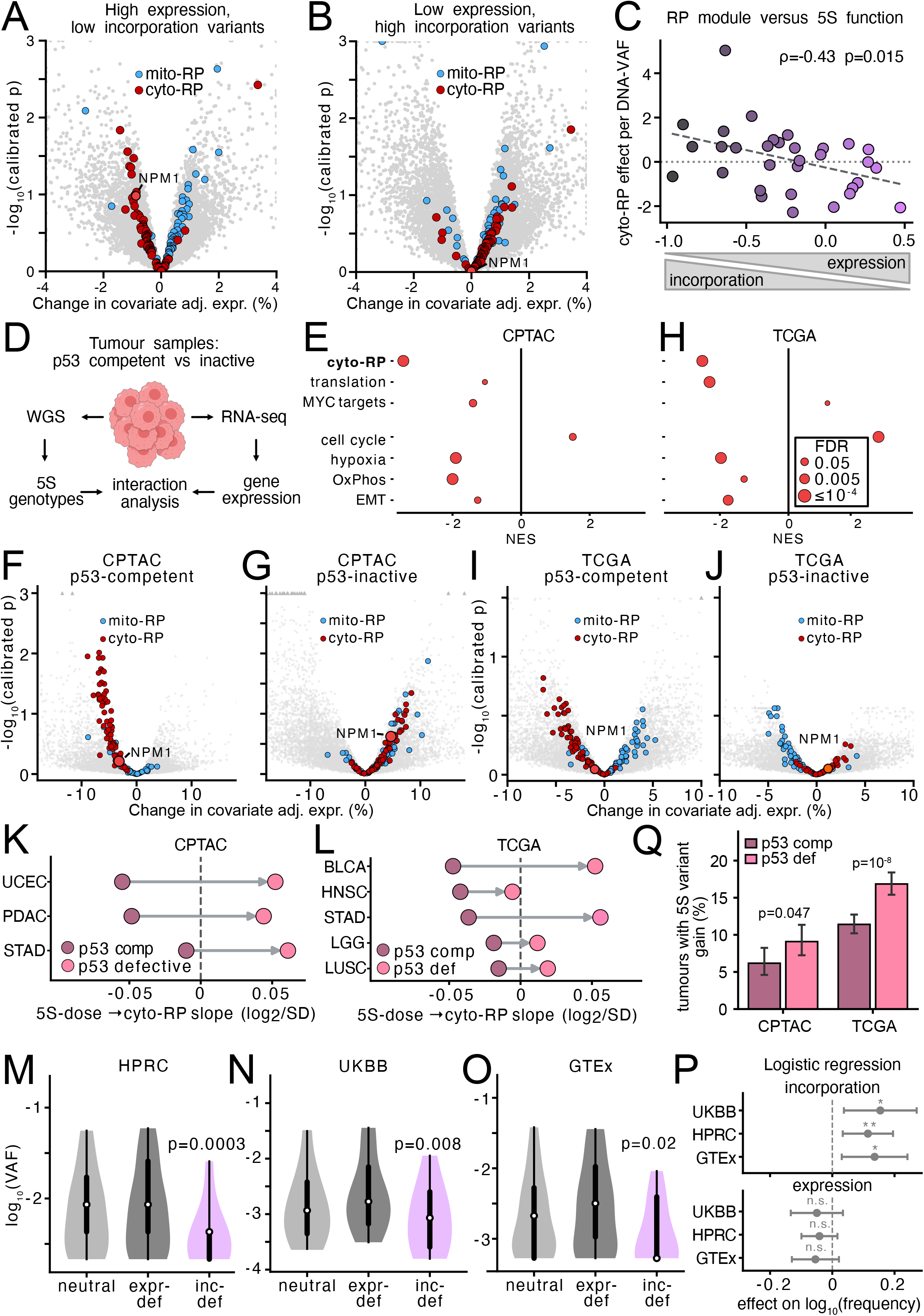
Incorporation-defective 5S rRNA variants are purified from human populations and repress ribosomal-protein genes through a p53-dependent program in cancer. **(A, B)** GTEx trans-effect of genomic 5S variant load, stratified by variant function: volcano of change in covariate-adjusted expression (%) versus - log_10_(permutation-calibrated p) for **(A)** high-expression/low-incorporation and **(B)** low-expression/high-incorporation variants; cyto-RP (red), mito-RP (blue), *NPM1* marked. Incorporation-defective load downregulates cyto-RP (83% of genes); functional load does not. **(C)** Per-variant cyto-RP module effect (per DNA-VAF) versus position on the incorporation–expression axis (Spearman ρ=-0.43, p=0.015; dashed line, fit). **(D)** Cancer-analysis design: tumour WGS→5S genotypes and matched RNA-seq→expression combined in a p53-stratified interaction analysis (CPTAC, TCGA). **(E)** GSEA of the transcriptome against genomic 5S dose in the p53-competent compartment for CPTAC; NES per pathway (blue, DNA; red, RNA in CPTAC), dot size ∝ log(FDR); ribosome/translation programs coordinately repressed. **(F, G)** CPTAC λ-calibrated trans-effect volcanoes (change in covariate-adjusted expression % versus -log_10_ calibrated p) for the **(F)** p53-competent and **(G)** p53-inactive fractions; cyto-RP (red), mito-RP (blue), *NPM1* marked; cyto-RP repressed in competent (GSEA FDR q<0.001), unaffected/reversed in inactive. **(H)** As in **(E)** for TCGA. **(I, J)** As in **(F, G)** for TCGA. **(K, L)** Per-type cyto-RP module slope (log2 per SD of 5S DNA dose) in p53-wild-type (blue) and p53-mutant (red) fractions for RP-down types with a testable mutant subgroup (n_mut>15) in **(K)** CPTAC (UCEC, PDAC, STAD) and **(L)** TCGA (BLCA, HNSC, STAD, LGG, LUSC); the association is uniformly weaker or reversed in the mutant fraction. **(M–O)** Per-variant carrier frequency [log_10_(VAF)] for neutral, expression-defective (<50% expression) and incorporation-defective (<50% incorporation) 5S gene variants in **(M)** HPRC, **(N)** UKBB and **(O)** GTEx; incorporation-defective variants exhibit lower carrier frequencies (violins; white dot, median; box, IQR; two-sided p = 3×10⁻⁴, 8×10⁻³, 2×10⁻²). **(P)** Multivariable regression of carrier frequency [log10(frequency) ∼ incorporation + expression + is_transition + is_CpG]; forest of the incorporation (top) and expression (bottom) coefficients per cohort (point, estimate; whiskers, 95% CI). Incorporation-defective variants remain depleted (∼33% less frequent; pooled β=+0.23, p=2×10⁻⁸; *) whereas expression is not (β=-0.08, n.s.). **(Q)** Fraction of tumours (%) carrying ≥1 significant somatic 5S gene-body gain (|ΔVAF| ≥ 1%, tumour vs matched blood-normal WGS), in CPTAC and TCGA, split by p53-competent (grey) vs p53-inactive (red; *TP53* mutation/deletion, *MDM2* or *MDM4* amplification, or *CDKN2A*/*ARF* deletion, plus HPV⁺ for CPTAC). Error bars, Wilson 95% CI (tumour-level); p, two-sided Fisher exact.

### Incorporation defective 5S variants act through the p53 pathway

We next asked whether this transcriptional signature, repression of the cytosolic ribosomal-protein (RP) regulon, is mediated by p53. The 5S RNP–p53 axis predicts that it should: variant 5S rRNA that is transcribed but cannot be incorporated into the ribosome accumulates as free 5S RNP, which stabilises p53^6–8^. We therefore tested whether p53 is required for the RP repression we observe. We analysed matched genetic and gene expression data from the Clinical Proteomic Tumour Analysis Consortium (CPTAC). We extracted 5S rDNA genotypes from WGS of 1426 tumours and defined each tumour’s genomic 5S variant dose as the summed WGS variant allele fraction across the 5S gene body. We then analyzed RNA sequencing data to understand 5S variation associated gene expression changes (**Figure 5D**). Tumours were classified by p53 status, incorporating non-genetic inactivation (*MDM2* or *MDM4* amplification, *CDKN2A*/*ARF* deletion) alongside *TP53* mutation. Among the five tumour types with a substantial p53 wild-type fraction (above 20%: CCRCC, UCEC, STAD, PDAC, LUAD), four showed the genomic 5S dose to RP-down signature in their wild-type compartment (**Figure 5E, S5C**) (CCRCC, UCEC, STAD and PDAC but not LUAD), validating this result from the GTEx cohort (**Figure 3J**). The remaining types (GBM, HNSC, LSCC) each had fewer than 20% wild-type samples, limiting power to detect a wild-type specific signature. In the four responsive types we asked whether p53 loss abrogates this relationship. Indeed, while the p53 competent fraction of these tumours showed robust and significant (FDR q < 0.001) repression of the cyto-RP module **(Figure 5F**), the p53-inactive fraction showed no significant relationship (**Figure 5G**). This suggested that the repression of ribosomal mRNAs was mediated by accumulation of the 5S RNP and activation of p53. Next, we turned to The Cancer Genome Atlas (TCGA) as an independent validation cohort. Seven tumour types showed a directional genomic 5S dose to RP-down association (BLCA, STAD, HNSC, LGG, KIRC, LUSC, PCPG, **Figure 5H**), and in the types with a testable p53 mutant subgroup the association was again weaker or reversed in the p53-mutant fraction, reproducing both the direction and magnitude seen in CPTAC (**Figure 5I, J**). Notably, this attenuation was present in every testable tumour type. Every RP-down tumour type with a sufficient p53-mutant subgroup (n >15), three in CPTAC (PDAC, UCEC, STAD) and five in TCGA (BLCA, STAD, HNSC, LGG, LUSC), showed a reduced or abrogated 5S dose to RP-down association in its mutant fraction (**Figure 5K, L, Table S9**). In summary, this consistency across histologically and genetically independent cancer types, in two separate cohorts, indicates that deleterious 5S rDNA alleles repress ribosome components in a p53-dependent manner.

### Incorporation defective 5S variants are purged from the human population

We aimed to understand how these functional impacts of 5S biology interacted with our population genetics findings. Thus, we investigated whether the frequency of 5S variants was correlated with functional alterations in the three cohorts (HPRC, UKBB and GTEx). Strikingly, incorporation defective (<50% incorporation) variants (**Figure 4G**) were significantly less frequent than neutral or expression defective (<50% expression) variants (**Figure 4E**) in all three cohorts (**Figure 5M-O**). We aimed to confirm that this finding was robust and specific. To this end, we built a regression model (log_10_(frequency) ∼ incorporation + expression + is_transition + is_CpG) to measure whether this trend held up after correcting for substitution and CpG frequency. Incorporation deficient variants remained significantly depleted in all three cohorts (**Figure 5P, Table S10**) and were on average 33% less frequent than other variants. This constraint was specific to the incorporation step: in a joint model the incorporation coefficient was strongly positive (pooled β=+0.23, p=2×10⁻⁸), whereas expression-defective variants showed no comparable depletion (expression β=-0.08, ns), confirming that selection targets assembly, not transcription or stability. Both cancer cohorts showed the same depletion of incorporation defective variants in germline (whole blood) WGS (**Figure S5E-G**). To test whether p53 shapes selection at the 5S rDNA locus, we examined somatic 5S variation in tumour genomes (CPTAC and TCGA). In both cohorts, somatic gains at the 5S locus were ∼50% more frequent in functionally p53-deficient than in p53-competent tumours (**Figure 5Q, Table S11**), consistent with p53 surveillance normally restraining amplification of 5S variants. In summary, incorporation defective 5S rDNA variants are purged from the human population driven by the p53 pathway.

## DISCUSSION

Human ribosomal DNA has been the last major functional locus inaccessible to genome-wide analysis, excluded by its repetitiveness until telomere-to-telomere assembly made it tractable^2^. By combining ∼600 T2T-resolved 5S haplotypes, variant calling in ∼490 000 UK Biobank participants, tissue-resolved GTEx expression, a saturation-mutagenesis map of 5S function, and matched genotype-expression data in two cancer cohorts, we trace human 5S rDNA variation from its evolutionary origin through its expression, and ribosome incorporation to its purging from the population. This 5S variation is not inert; it is expressed, perturbs ribosome assembly, is read out by the 5S RNP-p53 axis, and is under purifying selection. Together with parallel work on 45S copy number and 28S sequence variation^4,12,13,21,26^, this establishes the rDNA arrays as a functionally consequential, multi-dimensional source of human variation.

### Widespread but constrained 5S heterogeneity

Tandem rDNA is the textbook case of concerted evolution; unequal crossover and gene conversion homogenize copies within a species^27,28^. Homogenization is efficient but incomplete, and per-copy heterogeneity has been recognized since Arnheim and Southern (1977)^29^. Recent long-read surveys converted this expectation into catalogued 45S variants and proposed the existence of heritable expansion-segment “ribosome subtypes”^3,13,30^. We extend this to the unexamined 5S locus and beyond cataloguing and phenotype association to a mechanistic chain: variant expression, incorporation, and p53-dependent consequences. The contrast between the two arrays is instructive. Variation in the surface-exposed expansion segments of the 18S and 28S rRNAs have been proposed to generate specialized ribosomes with distinct functions^10,11^. Whether such variants are adaptive or merely tolerated remains largely untested. The 120-nucleotide 5S rRNA behaves differently: it is under strong constraint, partially protected from gene conversion, and most tested variants reduce rather than reprogram function. Human 5S variation is therefore a constrained axis of ribosome heterogeneity in which the dominant fate of a variant is diminished assembly, a distinction that, as below, has direct consequences for selection. Why expressed variants are enriched in brain, mirroring 45S variants in mouse brain, needs to be explored in the context of lineage-specific deployment or relaxed quality control^3^.

### A surveillance-coupled selection filter and disease alleles in repeats

Beyond its structural role in the ribosome, 5S rRNA with uL18 and uL5 forms the 5S RNP, which upon impaired ribosome biogenesis binds and inhibits MDM2 to stabilize p53^6–9^. Our functional map localizes incorporation-defective variants to the first ∼45 nucleotides, coincident with the uL5/uL18 footprint – the interface that decides whether 5S is assembled or diverted into the free, p53-activating particle. This explains the selection signature: incorporation-defective variants are depleted ∼33% across three independent cohorts, and the constraint is specific to assembly, not transcription or stability. Thus a variant that is never expressed is invisible and near-neutral, whereas one expressed but not incorporated generates free 5S RNP, activates p53, and carries a fitness cost. Two independent readouts in human tumours point in the same direction: genomic load of incorporation-defective variants represses the cytosolic ribosomal-protein regulon specifically in the p53-competent compartment across multiple CPTAC and TCGA cancer types, and once this surveillance brake is released, p53-deficient tumours accumulate more somatic 5S gains. Human 5S variation is thus a natural, dose-graded perturbation of the 5S RNP-p53 axis. This raises the possibility that high-VAF incorporation-defective 5S alleles or extreme copy number states underlie phenotypes that have escaped diagnosis because the locus was unsearchable; our dose-dependent associations (e.g., obsessive-compulsive disorder, systemic sclerosis) are hypotheses for targeted resequencing. More broadly, this joins the effort to bring the repetitive genome into human genetics alongside VNTRs and other tandem repeats^31^, and our T2T-calibrated short-read caller offers a template for other collapsed arrays.

### Limitations of study

The edge pattern is correlative for transcription; and the functional map derives from over-expression in one cell line.

## MATERIALS AND METHODS

## KEY RESOURCES

### Genomic datasets

Reference assemblies. The T2T-CHM13v2.0 assembly (GenBank GCA_009914755.4) was used as the primary reference, defining the 5S rDNA array locus on chromosome

1. The Genome in a Bottle HG002 reference assembly was obtained from the GIAB Consortium (NCBI BioProject PRJNA200694).

Pangenome assemblies. Human Pangenome Reference Consortium (HPRC) phased diploid assemblies (Year 1 and Release 2; NCBI BioProject PRJNA730823; data portal https://github.com/human-pangenomics and https://s3-us-west-2.amazonaws.com/human-pangenomics) provided the main assembly variant catalogue, including CHM13 and GIAB HG002 (235 donors / 466 haplotypes). Corresponding long-read data for HPRC donors, Oxford Nanopore and PacBio HiFi, reside within the same BioProject (PRJNA730823). As an independent East-Asian population set, we used Chinese Pangenome Consortium (CPC) assemblies: CPC Phase I^32^ (58 individuals), obtained as finished per-sample assembly FASTA files from the Human Population Omics Group POG portal (https://pog.fudan.edu.cn/cpc); together with the complete diploid genomes T2T-YAO (GWH assemblies GWHDQZJ00000000, GWHDOOG00000000 and GWHDQZI00000000; raw reads GSA-Human HRA004987) and CN1 (GWH assembly GWHCBHP00000000, maternal GWHCBHM00000000 and paternal GWHCBHQ00000000; BioProject PRJCA016397). These were integrated as an independent population set by lossless pangenome-graph (W-line) extraction, totalling 60 genomes / 120 haplotypes.

Population and clinical cohorts. UK Biobank whole-genome sequences (∼490 000 participants) and linked ICD-10 diagnoses were accessed under application (UKBB APPLICATION ID 98772). GTEx v9 whole-genome sequencing and v11 bulk RNA-seq from matched donors were accessed through dbGaP (study phs000424) under authorized-access project #43672.

Cancer cohorts. The Cancer Genome Atlas (TCGA; n = 11,162, phs000178) and Clinical Proteomic Tumor Analysis Consortium (CPTAC; n = 2,024, with 5S rDNA analysed on 1,426 whole-genome tumour samples, phs001287) sequence data were accessed via the NIH Genomic Data Commons under dbGaP authorized-access project #44042 (TCGA request #157798-1; CPTAC request #157800-1; CPTAC-3 sub-cohort).

### Software

Sequence search/alignment: BLAST+, minimap2, MAFFT. General analysis: Python (NumPy, pandas, SciPy, statsmodels), SQLite.

## METHODS

### 5S rDNA array reconstruction and variant catalog from genome assemblies

We located the 5S rDNA array in each phased assembly by homology search against the 5S repeat consensus and extracted all repeat copies, including those split across assembly contigs, ordering copies along each array in their resolved 5′ to 3′ orientation. Each copy was aligned to a population consensus repeat unit (**Table S5**) and variants (single-nucleotide variants and indels) were called relative to that consensus. Copies were annotated by region (5S gene, antisense Alu SINE, non-transcribed spacer) using the repeat-unit feature map. Per-haplotype copy number, per-variant prevalence (copies and donors), and the substitution spectrum (normalized to eligible bases) were computed from this catalog. The primary assembly catalog comprised CHM13, GIAB HG002, and HPRC genomes (235 donors, 466 haplotypes) and East-Asian assemblies from the Chinese Pangenome Consortium (CPC Phase I, T2T-YAO, and CN1; 60 genomes, 120 haplotypes).

### Repeat-unit annotation

The 2.2 kb consensus repeat unit was annotated by combining homology search, microsatellite/low-complexity detection, and the canonical POLIII promoter architecture, yielding the 5S rRNA gene and its internal control region (Box A, intermediate element, Box C), the polymerase III terminator, an antisense Alu SINE, and the flanking GC-rich spacers and microsatellites.

### Gene-conversion and clustering analyses

Within-array spatial clustering of variants was quantified per haplotype with a runs-based clustering index. For a variant carried by k of an array’s n linearly ordered interior copies, we counted the number of contiguous carrier runs (R) and the corresponding adjacencies (A = k - R), and referenced them to the Wald–Wolfowitz exact null for k carriers placed at random among n ordered copies (expected adjacencies A_exp = k(k - 1)/n). The clustering index, (A_obs - A_exp)/(A_max - A_exp) where A_max = k - 1, is 0 when carriers are randomly placed and 1 when they are fully contiguous; per-site values were aggregated to a per-haplotype index and tested against the null across haplotypes (Wilcoxon signed-rank). Gene conversion was assessed from (i) the directional substitution spectrum as a function of variant tract size and carrier VAF (GC-biased gene conversion manifesting as excess A/T→G/C), and (ii) the rate and contiguity of large multi-copy variant tracts, compared across regions (5S gene, Alu, NTS) with bootstrap confidence intervals.

### Content-adjusted site-frequency spectrum by region

Bi-allelic SNVs from the assembly variant catalog (array-member copies, unmasked) were assigned a population frequency as the number of the 589 haplotypes carrying each (position, alternative-allele) variant, and classified as rare (<=1% of haplotypes, i.e. <=6) or common (>5%, i.e. >29). Each repeat-unit position was assigned a mutational context (A, T, non-CpG C, non-CpG G, CpG C, or CpG G) from the population consensus sequence, and a per-context expected rate was estimated as the total rare (or common) alleles at positions of that context divided by (number of such positions x 589). For each haplotype and region (5S gene, 630-749; Alu SINE, 787-1,066; other NTS, the remainder), a content-adjusted density was computed as the observed rare (or common) allele count divided by the region’s expected count (summed per-context rates over the region’s positions). Regions were compared by paired Wilcoxon signed-rank test across haplotypes.

### Short-read 5S rDNA variant calling and specificity analysis

Because the tandem 5S array is collapsed to an artificial 17-copy unit in the hg38 reference, short-reads spanning the locus were mapped to one 5S reference repeat unit, so that reads from all array copies in an individual contribute to a single per-position pileup. Per-variant within-person variant-allele fractions (VAF) were estimated from this pileup and reflect the fraction of an individual’s array copies carrying a variant; because a variant present in only a few of the many copies appears at correspondingly low VAF, a low calling threshold is required. To define the set of sequences that could in principle cross-map with the array gene, we searched the 120 bp consensus 5S gene against the two references that the short-read data are (GRCh38) or could be (CHM13) aligned to: the GRCh38 full analysis set and the CHM13v2.0 T2T assembly. Searches used NCBI BLAST+ blastn (v2.16.0) with word_size 7 and e-value < 1e-3, and hits were retained as near-full-gene dispersed loci if they covered at least 90 bp of the gene and lay outside the chr1q42 array. This yielded ∼339 dispersed loci in GRCh38 and ∼337 in CHM13, present on every chromosome; 26 were >=95% and 5 were >=99% identical to the array gene at the gene level. To measure specificity of alignment, we used the CHM13v2.0 assembly as ground truth and simulated reads whose true genomic origin is known. From the region we actually genotype, we took each of the 127 transcribed gene copies of CHM13’s real chr1q42 array (gene plus 250 bp on each side) and, separately, all 337 CHM13 dispersed pseudogene loci with their native flanks (plus 250 bp). Uppercased sequences were used to generate 150 bp paired-end reads with wgsim (htslib v1.23; -d 320 -s 20 -e 0.002, no additional simulated polymorphism), yielding ∼400 000 reads per origin, and reads were labelled by origin. All reads were aligned to GRCh38 with bwa mem (v0.7.19) and each primary alignment was classified by position as landing in the array window (chr1:228,609,921-228,646,357, together with the equivalent collapsed-array positions), on a dispersed 5S locus (within 250 bp of a catalogued locus), on other unique sequence, or unmapped. Because true 5S variants change the gene sequence, we confirmed that specificity does not depend on the gene matching the reference. We simulated 150 bp paired-end reads from both strands of the array window and of all dispersed loci, introduced a fixed number of substitutions per read (0, 1, 2, 3, 5, 8 or 12 substitutions at random positions), realigned to GRCh38, and measured leakage in both directions: array reads mis-mapping onto a dispersed pseudogene (loss of true signal) and pseudogene reads mis-mapping into the array window (contamination).

### Variant-calling optimization and benchmarking

The short-read 5S caller was developed and benchmarked against a ground-truth variant set built from the matched HPRC genome assemblies in donors that also had long-read data (n=41 HPRC year 1 donors). Short-read calls were scored as true positives, false positives, or false negatives relative to this assembly truth set. The calling threshold of 0.3% was set by sweeping the calling VAF threshold and choosing the operating point that maximized the balance of sensitivity and precision (F1), with performance further stratified by variant copy number (sensitivity) and by VAF (precision). A fraction of false positives are genuine variants the assembly missed; these were independently confirmed with PacBio HiFi long-reads (“rescue”). To establish that HiFi-confirmed calls reflect shared biological signal rather than coincidental cross-platform agreement, their overlap was tested against a permutation null that preserves each platform’s empirical error positions. At the chosen operating point (0.3% VAF), the caller reached a sensitivity of 83% and precision of 69%; boundary positions of the repeat unit which had lower SR coverage were excluded from calling (Figure 2A–C; Figure S2A–B).

### UKbiobank 5S array read extraction

Target regions corresponding to the 17 copies of the 5S rDNA gene were defined on the GRCh38 assembly raging from chr1:228,610,069 to chr1:228,646,359. WGS CRAM files aligned to the GRCh38 reference genome using DRAGEN from the UK Biobank 500K release were retrieved from data field 24048 for 490,086 participants with available data. For each participant, reads overlapping the region of interest were extracted using samtools view with -M and -L options. All analyses were performed on the DNAnexus Research Analysis Platform. This research has been conducted using the UK Biobank Resource under application number 98772.

### UK Biobank and GTEx short-read 5S rDNA genotyping

Whole-genome short-reads from UK Biobank (UKBB) GRCh38 alignments were extracted from the 5S rDNA locus on chromosome 1 (chr1:228,533,997–229,217,052) and genotyped as above, pooling reads per individual to estimate per-variant VAF. Per-position carrier counts then defined the population variation landscape across ∼490 000 participants. GTEx donors (n=943) were genotyped identically from their GRCh38 whole-genome alignments: reads over the 5S rDNA cluster (chr1:228,605,000–228,650,000) were extracted, mapped to the 5S reference unit, and pooled per donor to estimate per-variant DNA VAF (mpileup with mapping- and base-quality filters; minimum alt-depth and VAF as above). These per-donor DNA VAFs provide the genotype paired with RNA expression in the GTEx analyses below.

### Phenome-wide association and dosage analysis

We performed a phenome-wide association study (PheWAS) relating each 5S rDNA variant to the UK Biobank ICD-10 diagnosis catalogue across the ∼430 000 participants with linked ICD-10 records, age, sex, and genetic principal components. For every variant–diagnosis pair we fit a covariate-adjusted logistic regression of diagnosis (case/control) on the variant predictor and covariates, evaluating ICD-10 outcomes: diagnosis ∼ variant dosage + age + sex + PC1–PC10, where the covariates were age, sex, and the first ten genetic principal components (controlling for ancestry and population structure). The predictor was the quantitative estimated variant copy dosage per individual, the within-person VAF scaled to the diploid array copy, so that the model tests dose-dependent effects across the full range of within-person allele burden. Models were fit by maximum likelihood (iteratively reweighted least squares, with a BFGS fallback for non-convergence), and significance was taken as the Wald test of the variant-dosage coefficient, with odds ratios and 95% confidence intervals reported. To restrict testing to well-powered, reliably genotyped variants we required a confident VAF ≥ 0.3%, at least 50 carriers per variant, and a minimum number of exposed cases, and used a 2×2 pre-screen to skip variants with no nominal signal before fitting. Multiple testing was controlled by the Benjamini–Hochberg false-discovery rate (FDR < 0.05) applied within each ICD-10 block. Positional enrichment of FDR-significant associations in the 5S gene relative to the surrounding NTS was assessed both by Fisher’s exact test and, as the primary calibration given the modest number of hits, by a one-sided label-permutation test (20 000 permutations of the significant-hit labels across variants). For selected lead associations, dose–response was characterized as the odds ratio versus non-carriers across bins of estimated variant copy number (logistic regression).

### 5S rRNA variant expression in GTEx cohort

For matched GTEx donors (n=943), bulk RNA-seq reads over the 5S gene region were pooled per donor to estimate per-variant RNA VAF, paired with the donor’s DNA VAF from whole-genome sequencing, which is described above. Variant expression was assessed threshold-free by a carrier-versus-non-carrier rank statistic (Mann–Whitney U, expressed as the AUC) per variant: of 160 testable variants, 52 were classified as expressed (carrier-high; AUC indicating higher RNA-VAF in carriers than non-carriers, rank-skew FDR < 0.10) and none showed silencing. Donor-level prevalence of variant expression was estimated against a per-locus chance-null: a donor was called as expressing a given variant when its pooled RNA-VAF exceeded the non-carrier 99th percentile at that locus (alt-depth ≥ 3; ∼1% false-positive rate by construction). Across the 52 expressed loci, 15% of the 943 donors expressed ≥1 variant against a chance-null of 5%, giving ∼10% genotype-attributable expression. The DNA-versus-RNA VAF relationship (n=757 loci; RNA-VAF ∼6-fold below DNA-VAF) defined locus-specific transcription efficiencies.

### Transcriptome consequences of variant 5S carriage and expression

Genome-wide differential expression was modeled as a continuous function of 5S-variant dosage. For each of 943 donors we computed two aggregate, standardized dosage metrics over all 5S gene-region variants: a DNA dosage, the summed genomic variant-allele fraction across carried variants (from WGS), and an RNA dosage, the summed RNA-seq variant-allele fraction in excess of the matched non-carrier background. Within each of 37 tissues (19,244 samples in total) we fit a negative-binomial generalized linear model (DESeq2) of the form expression ∼ RIN + ischemic time + five genotype principal components + sex + Hardy death classification + sequencing batch + dosage, and tested the standardized dosage coefficient by Wald test; RNA and DNA dosage were assessed both marginally and together in a joint two-predictor model. Unmodeled technical structure was absorbed by the explicit sequencing-batch term. Per-gene effects were combined across tissues by fixed-effect inverse-variance meta-analysis (weighting each tissue by 1/SE^2), yielding a cross-tissue effect size, a Benjamini-Hochberg FDR, and a per-gene directional-consistency score. Pathway-level effects were summarized by gene-set enrichment analysis (GSEA, gseapy prerank; MSigDB Hallmark and Reactome plus custom cytosolic and mitochondrial ribosomal-protein sets) on the meta-analytic gene ranking. The cytosolic translation/ribosome program was the top-ranked of 1,834 gene sets in the RNA-dosage model (normalized enrichment score -3.8, FDR approximately 1×10^-3) and was coordinately downregulated with increasing variant-5S expression. Robustness was further assessed by restricting to genetically European donors and by the joint DNA/RNA model.

### Cloning of 5S rRNA plasmid and validation of overexpression

A ∼600bp repeat region of the human 5S rDNA repeat was amplified (For GGTGTAGGTGGGCGGTAAAG, Rev GATGTGGTGGAAGCTCGGG) from genomic DNA via polymerase chain reaction (PCR) and cloned into pJET1.2. To verify the expression of the 5S rRNA, the plasmid was transiently transfected into HEK293T cells alongside a GFP control. For this, 1.2 x 10^6^ cells were seeded per well in 6 well plates one day prior to transfection. Cells were transfected with 3μg of the respective plasmid DNA using polyethyleneimine (PEI) and incubating overnight at 37°C, 95% humidity and 5% CO_2._ Transfection efficiency was determined to be more than 90% by GFP fluorescence microscopy before proceeding with TRIzol-based total RNA extraction according to the manufacturer’s instructions. RNA concentration and purity was determined using a NanoDrop 2000 spectrophotometer and RNA integrity was assessed via capillary electrophoresis (Agilent 2100 Bioanalyzer). To evaluate 5S rRNA overexpression after transient transfection, the abundance of 5S rRNA was quantified relative to 5.8S rRNA. Peak height corresponding to 5S rRNA was quantified relative to 5.8S rRNA using Agilent 2100 Bioanalyzer software and the 5S/5.8S rRNA ratio was calculated for each sample.

### Saturation mutagenesis and functional testing of human 5S rRNA

Then, a plasmid library containing single nucleotide variants of the transcribed region of 5S rDNA was generated using the SUNi mutagenesis approach^33,34^. For this, degenerate oligonucleotides were designed such that each nucleotide position within the 5S rDNA gene was substituted by all possible alternative nucleotides. Diversity of the plasmid library was evaluated by PCR and amplicon sequencing. Sequencing reads were then analyzed to determine variant representation and coverage of the expected SNVs. In the final library, 84% of possible SNVs were detected, distributed over a ∼32-fold frequency range. For functional testing, the plasmid library or a GFP encoding plasmid were transiently transfected into HEK293T cells using PEI. Cell lysates were prepared by first aspirating the growth medium and washing the cells once with freshly prepared wash buffer (1x PBS, 10mM MgCl_2_, 800μM cycloheximide). Excess buffer was removed by resting the plates in a tilted manner for 10 seconds, so that the excess liquid can collect at the bottom, before removing it with a pipette. Cells were lysed with lysis buffer (0,25M HEPES (KOH) pH 7.5, 50mM MgCl_2,_ 1M KCl, 5% (vol/vol) NP40, 1000μM CHX) by scraping the cells with a cell scraper and collecting the lysate with a pipette from the bottom of the plates. Lysates were then clarified by centrifuging at 20 000 rpm for 15min at 4°C. The resulting samples were fractionated by ultracentrifugation on a 10–50% sucrose gradient (20mM Tris pH 7.4, 20mM MgCl_2,_ 320mM KCl) at 40 000 rpm for 3 h at 4°C. Fractions corresponding to the low-density fraction (cytosol), 60S, 80S and polysomes were collected using the Biocomp gradient station and subsequently subjected to acid-phenol–chloroform based RNA extraction. Samples for deep sequencing were prepared according to the mim-tRNA-seq library preparation workflow^35^. This protocol was followed, except intermediate products of the different expected size we excised and PCR was performed in the presence of 0.5M betaine. The library was then subjected to deep-sequencing on an Illumina Nextseq 2000. Sequencing reads were demultiplexed, adapter trimmed with cutadapt and quality filtered. Reads were then aligned to a consensus sequence of 5S rDNA. Mismatches and coverage were counted over the positions and the GFP transfected samples were used to determine and subtract the local sequencing error rate. Corrected substitution rates were used as sample specific variant frequencies.

### Cross-cohort purifying-selection analysis

Each functionally assayed gene variant was assigned an expression score and a 60S-incorporation score from the saturation-mutagenesis assay, and classified for visualization as neutral, expression-defective, or incorporation-defective (defect <0.5 of wild-type). Population frequency was computed as a per-individual confident-carrier frequency (VAF ≥ 0.3%) in three independent cohorts, UK Biobank, HPRC assemblies (an individual scored as a carrier if ≥0.3% of its copies carried the variant), and GTEx.Depletion of incorporation- versus expression-defective variants was tested by Mann–Whitney U; specificity was confirmed with a per-cohort ordinary-least-squares regression of log10 carrier frequency on the incorporation and expression scores, in which the incorporation coefficient estimates the effect on frequency at fixed expression.

### Use of AI and large language models

Large language models (LLMs), including OpenAI ChatGPT and Anthropic Claude, were used as coding and writing assistants during this study to help draft, debug, and document analysis code, to organise the code for public release, and for language editing of the manuscript. No LLM was used to generate, process, or interpret primary data, or to derive the scientific conclusions; all analyses were designed by the authors, and all AI-assisted code and text were reviewed, tested, and validated by the authors. The authors take full responsibility for the integrity and content of the study, including all AI-assisted components.

### Quantification and statistical analyses

Statistical tests, sample sizes (n), and dispersion measures are reported in the corresponding figure legends. Group comparisons used Mann–Whitney U or Wilcoxon tests; correlations used Spearman or Pearson; enrichment used permutation and Fisher’s exact tests; and multi-predictor effects used ordinary-least-squares or logistic regression. Multiple comparisons were controlled by the Benjamini–Hochberg false-discovery rate or Bonferroni correction. Where many sequence copies derive from few donors, donors were treated as the unit of replication to avoid pseudoreplication. Analyses used Python (NumPy, pandas, SciPy, statsmodels).

## Data Availability

An interactive resource for exploring the 5S rDNA variation and functional data is available at https://genesintranslation.com/. Sequencing data generated in this study have been deposited in the NCBI Gene Expression Omnibus (GEO) under accession GSE339543. This study also analysed previously published and controlled-access datasets, HPRC assemblies and long-read data (BioProject PRJNA730823), Chinese Pangenome Consortium assemblies, UK Biobank (Application 98772), GTEx (dbGaP phs000424), and TCGA and CPTAC (dbGaP phs000178 and phs001287), available from their respective repositories under the accessions and data-access procedures listed in the Methods. Derived data supporting the findings are provided in the Supplementary Tables. All analysis code is available at https://github.com/Bohlenlab/5S-rRNA-surveillance and archived on Zenodo (DOI: 10.5281/zenodo.21743975). Plasmids and other reagents generated in this study are available from the lead author (Jonathan Bohlen) upon request.

https://zenodo.org/records/21743975

https://github.com/Bohlenlab/5S-rRNA-surveillance

https://genesintranslation.com/

## AVAILABILITY OF DATA, CODE AND MATERIALS

An interactive resource for exploring the 5S rDNA variation and functional data is available at https://genesintranslation.com/. Sequencing data generated in this study have been deposited in the NCBI Gene Expression Omnibus (GEO) under accession GSE339543. This study also analysed previously published and controlled-access datasets, HPRC assemblies and long-read data (BioProject PRJNA730823), Chinese Pangenome Consortium assemblies, UK Biobank (Application 98772), GTEx (dbGaP phs000424), and TCGA and CPTAC (dbGaP phs000178 and phs001287), available from their respective repositories under the accessions and data-access procedures listed in the Methods. Derived data supporting the findings are provided in the Supplementary Tables. All analysis code is available at https://github.com/Bohlenlab/5S-rRNA-surveillance and archived on Zenodo (DOI: 10.5281/zenodo.21743975). Plasmids and other reagents generated in this study are available from the corresponding author upon request.

## ACKNOWLEDGEMENTS AND FUNDING

We thank Evangelos Karousis for critical reading of the manuscript and constructive discussions. We thank Prof. Dr. Shuhua Xu for assistance with obtaining access to the Chinese pan genome consortium data. The Bohlen lab is supported by an Emmy-Noether Grant (DFG), funding from the TRR237 consortium (INST 269/1131-1, DFG), the Ludwig-Maximillians Universtiy (LMU), the Daimler Benz Foundation (Grant 32-01/25) and a Life Science Bridge Award from the Aventis Foundation. The work was supported by German Research Foundation (DFG) grants CRC237 369799452 (J.K.) and an Else-Kröner-Fresenius-Stiftung starting grant 2019_A70 (J.K.).

## AUTHOR CONTRIBUTIONS

L.S, J.B., I.B., and M.M. carried out experiments. C.C., Y.S., A.C. and J.B. were involved in data curation. L.S. and J.B. carried out data analysis. J.B. conceptualized and supervised the project. L.S. and J.B. wrote the initial manuscript, all authors were involved in review and editing of the manuscript.

## CONFLICT OF INTEREST

The authors declare that they have no conflict of interest.

## SUPPLEMENTAL FIGURE LEGENDS

**Figure S1.**
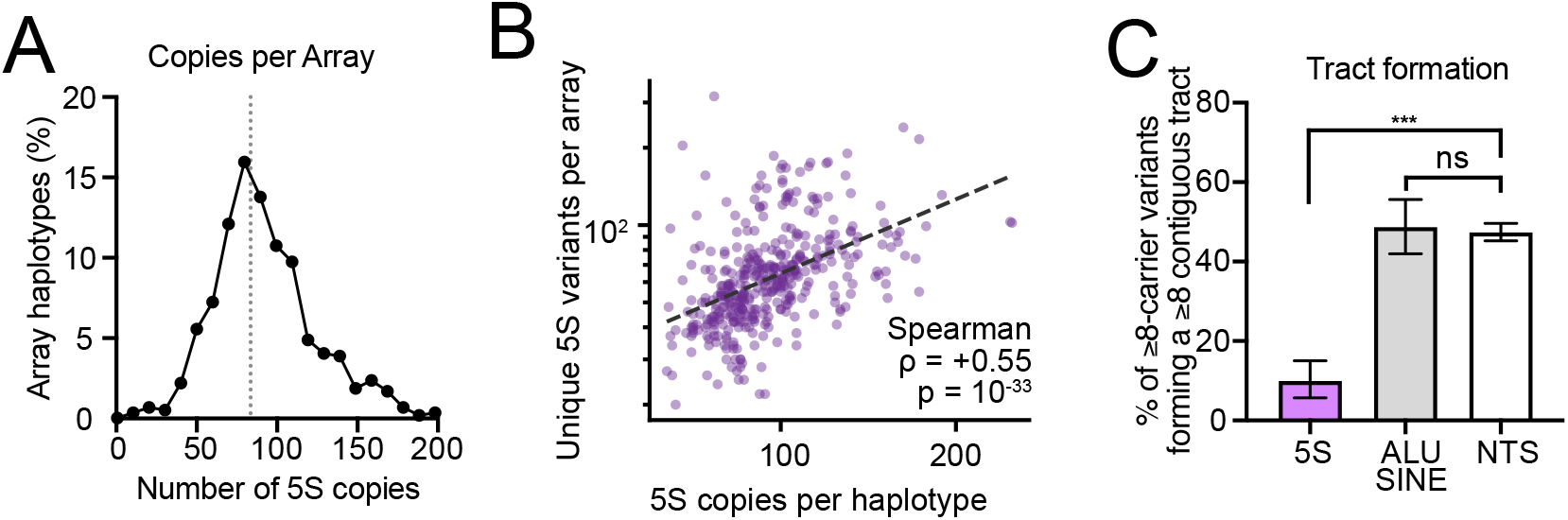
Characterisation of 5S repeat array architecture. Related to Figure 1. **(A)** Distribution of haploid 5S copy number across array haplotypes; red dashed line, median. **(B)** Each dot is one HPRC haplotype array (n = 410; resolved/single arrays, ≥20 copies). The number of unique 5S variants per array (distinct single-nucleotide variants across all copies, indels excluded; y-axis, log scale) is plotted against the array’s copy number (x-axis). Dashed line, log-linear fit. Copy number and unique-variant count are positively correlated (Spearman ρ = 0.55, p = 1×10⁻³³), consistent with variants accumulating in proportion to the number of template copies. **(C)** Fraction of ≥8-carrier variants that form a ≥8-copy contiguous tract, by region. Bars, mean ± bootstrap 95% CI; significance by logistic regression of tract formation on region with haplotype-clustered (cluster-robust) standard errors. *p<0.001; n.s., not significant.

**Figure S2.**
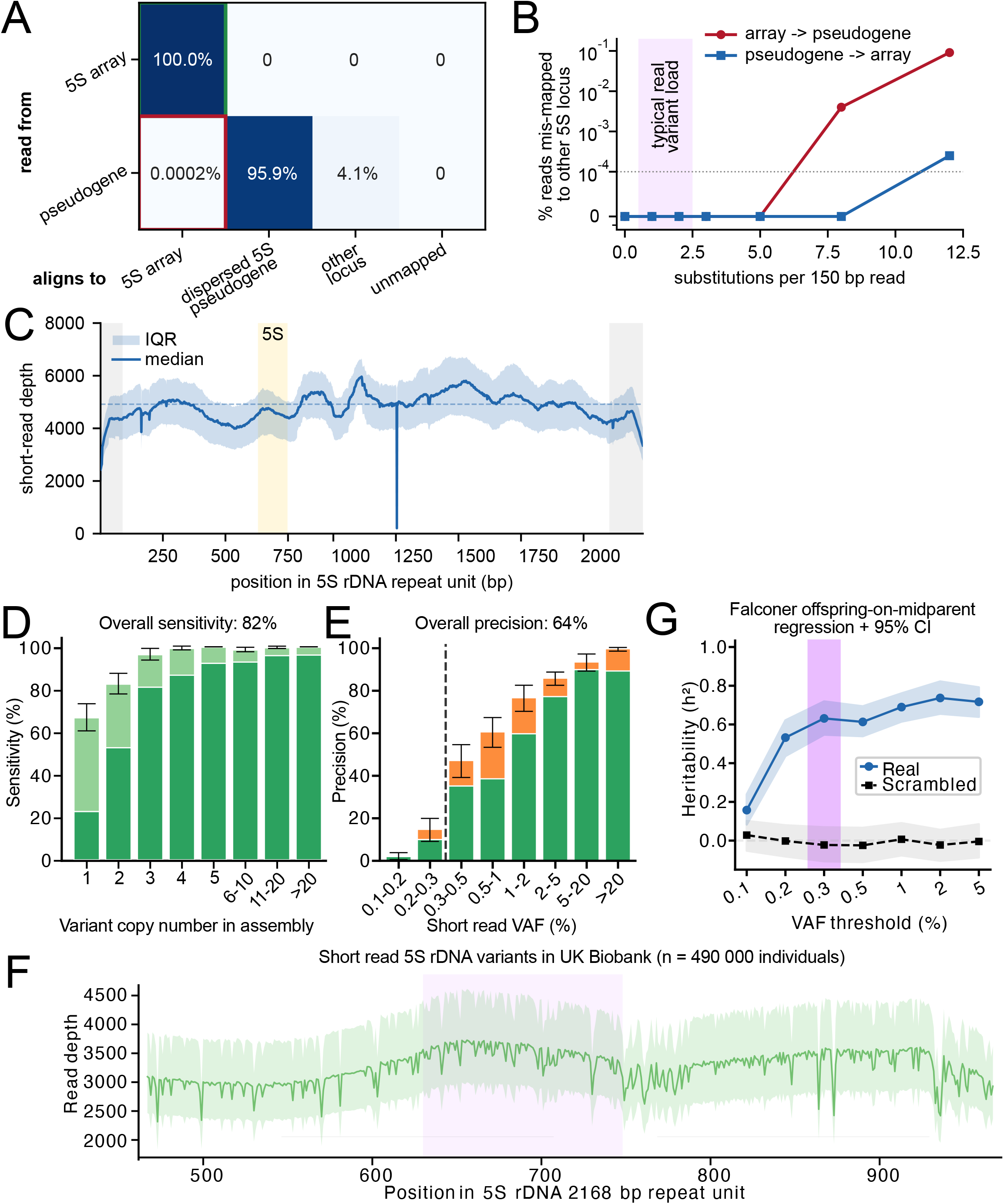
Validation of UK Biobank short-read 5S variant calling and heritability. Related to Figure 2. **(A)** Ground-truth read-level confusion matrix. 150 bp paired-end reads were simulated from sequences of known origin in the CHM13 T2T assembly, either the transcribed gene copies of the real chr1q42 5S array (127 copies, gene +/-250 bp; top row) or all 337 dispersed 5S pseudogene loci with their native flanks (bottom row), and aligned to GRCh38. Each cell gives the percentage of reads of a given origin (row) whose primary alignment lands at a given destination (column): the 5S array locus (the extraction window chr1:228,609,921-228,646,357 plus the equivalent collapsed-array positions), a dispersed 5S pseudogene, other unique sequence, or unmapped. **(B)** Cross-mapping is negligible and robust to sequence variation. Percentage of simulated 150 bp reads that mis-map to the other 5S locus type as a function of the number of substitutions introduced per read (both strands simulated), shown for array reads mis-mapping onto a dispersed pseudogene (red, loss of true signal) and pseudogene reads mis-mapping into the array window (blue, contamination); y-axis on a symmetric-log scale so exact zeros are shown on the baseline. **(C)** Short-read coverage of the 5S rDNA consensus is even across the repeat unit. Illumina WGS reads extracted from the chr1q42 5S array in HPRC Year-1 samples (n = 43) were re-aligned to a single 2168 bp population-consensus repeat unit, and read depth was computed per position (samtools depth). Grey, boundary positions excluded from variant calling. **(D)** Sensitivity of short-read calling versus variant copy number in the matched assembly (overall 82%); each bar is split into calls independently confirmed by HiFi long-reads (dark) versus SR-only detections lacking HiFi confirmation (light). Bars, mean ± 95% CI. **(E)** Precision versus short-read VAF (overall 64%); the dashed line marks the 0.3% calling threshold below which precision falls. Green: True positive assembly variants. Orange: additional fraction added by PacBio HiFi rescue. Bars, mean ± 95% CI. **(F)** Per-position read depth along the 5S repeat unit in UK Biobank (n≈490 000); line, mean; light band, 5th–95th percentile (across individuals); gene region shaded. **(G)** Heritability of 5S variant load by Falconer offspring-on-midparent regression versus VAF threshold, for real (blue) versus scrambled (black) pedigrees; h²≈0.6–0.7 above the calling threshold. Points, estimate; bands, 95% CI.

**Figure S3.**
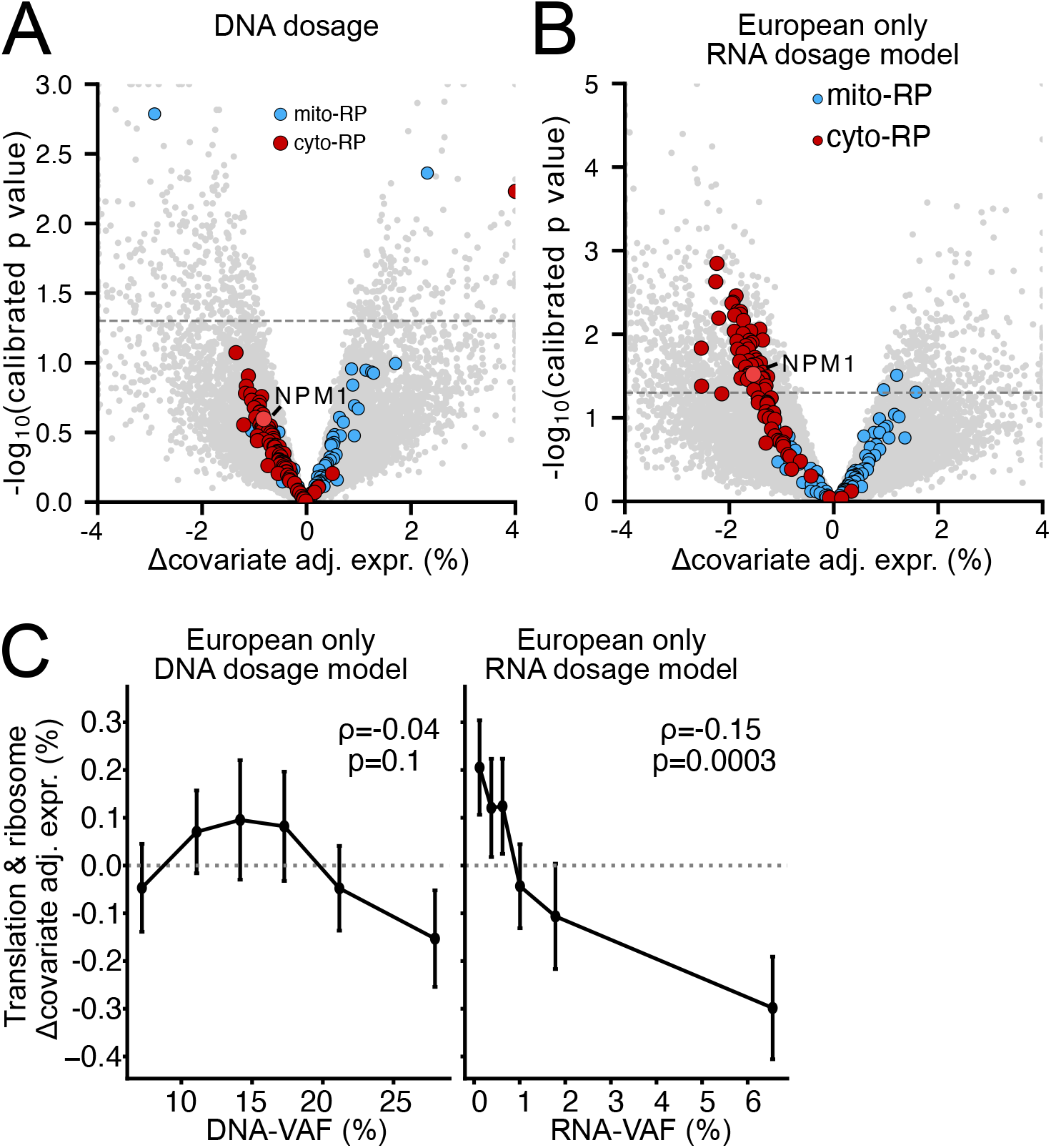
The 5S-dose trans-effect on ribosomal-protein genes is present across dosage metrics and ancestries. Related to Figure 3. **(A)** DNA-dosage-model trans-effect volcano (companion to the RNA-dosage model in Figure 3J): change in covariate-adjusted expression (%) versus -log_10_(calibrated p); cyto-RP (red) down, mito-RP (blue) unaffected, *NPM1* (orange). **(B)** RNA-dosage-model trans-effect volcano restricted to European-ancestry donors. **(C)** Dose-response of the translation/ribosome module in European-ancestry donors versus DNA-VAF (left; Spearman ρ=-0.04, p=0.1) and RNA-VAF (right; ρ=-0.15, p=0.0003).

**Figure S4.**
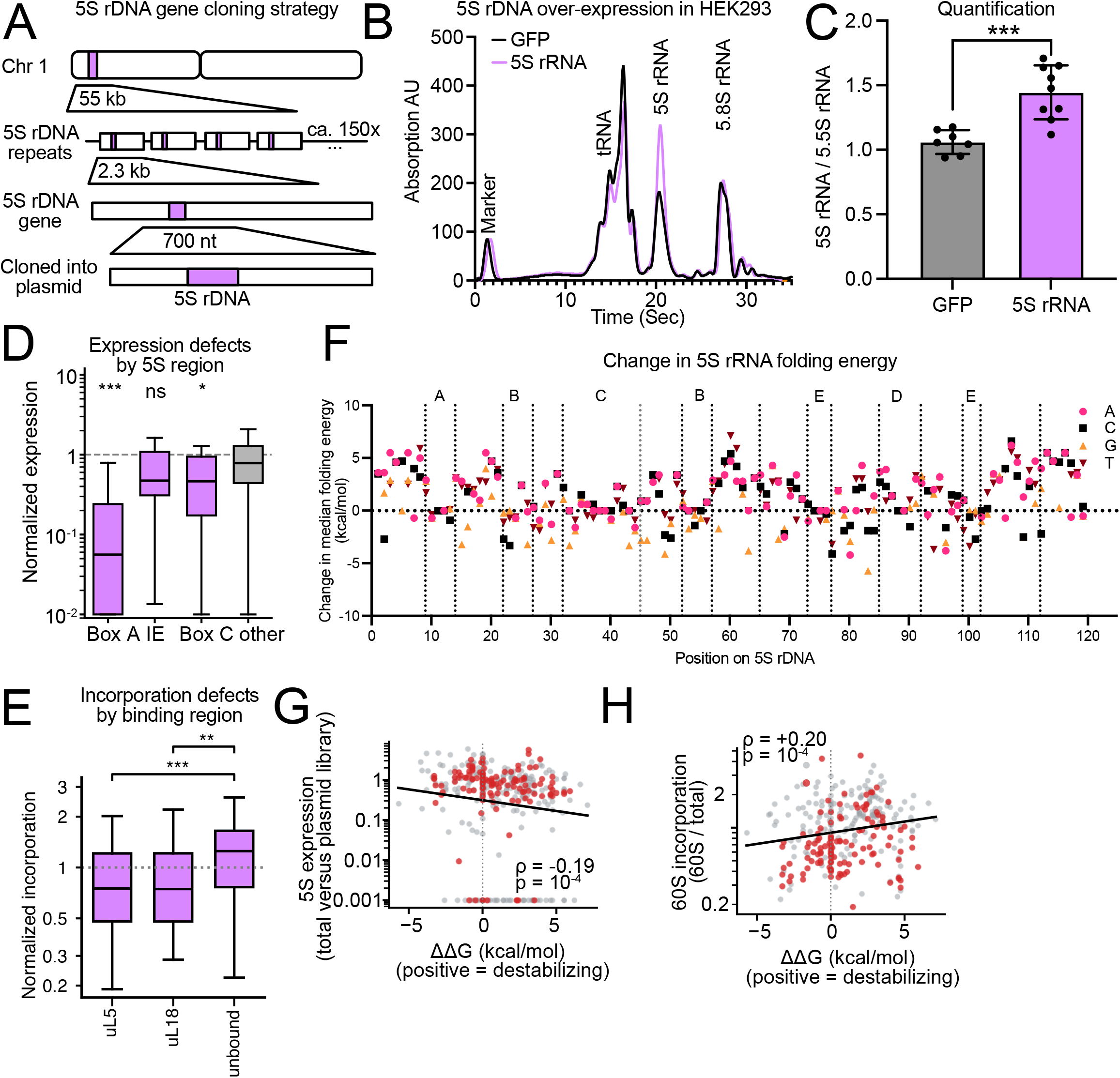
5S rDNA overexpression validates the expression readout. Related to Figure 4. **(A)** Cloning strategy: the genomic 5S rDNA gene (one unit of the ∼55 kb, ∼150-copy chromosome-1 array) is cloned into an expression plasmid. **(B)** Capillary-electrophoresis trace of total RNA from HEK293 cells transfected with the 5S rDNA construct (orange) versus a GFP control (black); the 5S rRNA peak is selectively increased (tRNA, 5S, and 5.8S rRNA peaks labeled). **(C)** Quantification of the 5S rRNA / 5.8S rRNA ratio (GFP vs 5S rDNA; bars, mean; points, replicates; p***<0.0005), confirming functional overexpression of the cloned 5S rDNA. **(D)** Normalized expression of variants grouped by 5S-gene region (Box A, IE, Box C, other); Box A and Box C variants reduce expression (***, ***), IE does not (n.s.) (box plots; median, IQR, whiskers). ***p<0.0005 **(E)** Incorporation (60S-vs-total variant frequency; as in Figure 4G) of variants at 5S positions contacting uL18 or uL5 in the 5S RNP versus non-contacting positions, using 8BGU for contact annotation. Variants at uL18 - and at uL5-contacting positions are both significantly more incorporation-defective than unbound positions (median 0.75 and 0.75 vs 1.25; Mann–Whitney p=3.1×10⁻⁷ and p=2.2×10⁻³), showing that incorporation defects concentrate at the 5S-RNP protein interface. **(F)** Predicted change in 5S rRNA folding energy (Δ median folding energy, kcal/mol) per position along the 5S gene (nucleotides A/C/G/T). **(G, H)** Predicted folding stability relates to 5S rRNA function. Per-variant change in folding free energy versus expression **(G)** and versus 60S incorporation **(H)**, for all assayed single-nucleotide variants of the 5S rRNA gene. Each point is one variant; variants in the first 45 nt (the uL5/uL18 contact footprint) are red, the remainder grey. Solid line, log-linear trend; dotted line, ΔΔG = 0 (wild-type stability).

**Figure S5.**
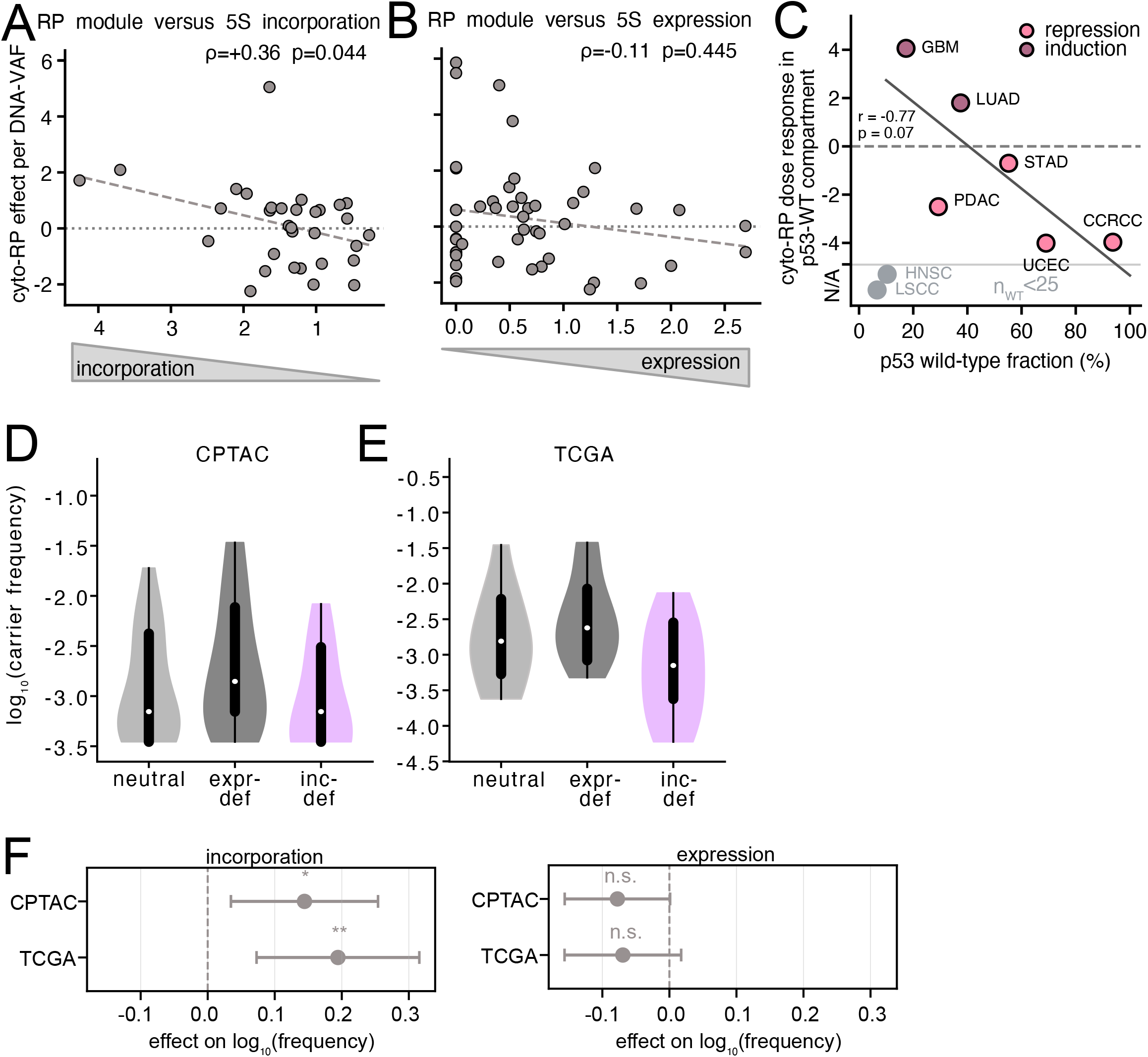
Incorporation, not expression, drives 5S-dose repression of ribosomal-protein genes; the p53-competent compartment scales with repression. Related to Figure 5. **(A)** Per-variant cyto-RP module effect (per DNA-VAF) versus the incorporation component (ρ=+0.36, p=0.044; dashed line, fit.) **(B)** As in **(A)** versus the expression component of the functional axis (Spearman ρ=-0.11, p=0.445). **(C)** Per CPTAC tumour type, cyto-RP dose-response (%/SD of 5S DNA dose) in the p53-wild-type compartment versus the p53-wild-type fraction (%), (Pearson r=−0.77, p=0.07; green, repression; orange, induction; solid line, fit); HNSC and LSCC (n_WT<25) shown at their wild-type fraction with no estimable effect (n/a) and excluded from the correlation. **(D, E)** Incorporation-defective 5S rRNA gene variants are depleted in the CPTAC and TCGA cohorts. Per-variant carrier frequency (confident carriers with VAF ≥ 0.3% divided by cohort donors; log scale) for neutral, expression-defective (<50% expression) and incorporation-defective (<50% incorporation) 5S gene variants in **(D)** CPTAC (N = 1,427) and **(E)** TCGA (N = 8,392) germline whole-genome sequences (whole blood), violins trimmed to the 10th–90th percentile; white dot, median; thick bar, IQR; thin line, 10th–90th percentile. **(F)** The frequency constraint is specific to incorporation, not expression, in both cancer germline cohorts. Multivariable regression of per-variant carrier frequency [log_10_(frequency) ∼ incorporation + expression + is_transition + is_CpG] in the CPTAC and TCGA germline cohorts; forest of the incorporation (top) and expression (bottom) coefficients per cohort (point, estimate; whiskers, 95% CI; dashed line, no effect).

## Notes

### Competing Interest Statement

The authors have declared no competing interest.

### Author Declarations

This study is a retrospective secondary analysis of existing, de-identified human datasets; no new participants were recruited and no identifiable data were accessed. All source cohorts were collected with informed consent under their own ethical approvals. UK Biobank holds Research Tissue Bank approval from the North West Multi-centre Research Ethics Committee (REC reference 21/NW/0157), which covers registered secondary analyses; its data were accessed under Application 98772. GTEx donors consented under the project's approved protocols, and individual-level whole-genome and RNA-sequencing data were obtained from dbGaP (accession phs000424) under authorized-access project 43672. CPTAC and TCGA tumour and matched-normal data were generated by contributing centres under local IRB approval with informed consent and accessed as controlled-access data via dbGaP / the NCI Genomic Data Commons (project 44042). Human Pangenome Reference Consortium and Chinese Pangenome Consortium assemblies, and the Fiber-seq datasets, are broadly consented resources released for open research use.

